# The Effect of Metformin Treatment on the Circulating Proteome

**DOI:** 10.1101/2024.06.07.24308435

**Authors:** Ben Connolly, Laura McCreight, Roderick C Slieker, Khaled F Bedair, Louise Donnelly, Juliette A de Klerk, JWJ Beulens, PM Elders, IMI-DIRECT, IMI-RHAPSODY, Göran Bergström, Mun-Guan Hong, Robert W. Koivula, Paul W. Franks, Leen ‘t Hart, Jochen M Schwenk, Anders Gummesson, Ewan R Pearson

## Abstract

**Objective:** Metformin is one of the most used drugs worldwide. However, its mechanism of action remains uncertain. Given the potential to reveal novel insights into the pleiotropic effects of metformin treatment, we aimed to undertake a comprehensive analysis of circulating proteins.

**Research Design and Methods:** We analysed 1195 proteins using the SomaLogic platform in 1175 participants, using cross- sectional data from the GoDARTS and DCS cohorts; 450 proteins using the Olink platform in 784 participants, using cross-sectional data from IMI-DIRECT; and combined longitudinal data from the IMPOCT, RAMP and S3WP-T2D cohorts with 372 proteins in 98 participants using the Olink platform. Finally, we performed systems level analysis on the longitudinal OLINK data to identify any possible relationships for the proteins changing concentration following metformin exposure.

**Results:** Overall, 97 proteins were associated with metformin exposure in at least one of the studies (P_adj_<0.05), and 10 proteins (EpCAM, SPINK1, t-PA, Gal-4, TFF3, TF, FAM3C, COL1A1, SELL, CD93) were associated in two independent studies. Four proteins, REG4, GDF15, REG1A, and OMD were consistently associated across all studies and platforms. Gene-set enrichment analysis revealed that the effect of metformin exposure was on intestinal tissues. In the longitudinal analysis 18% of proteins were significantly altered by metformin.

**Conclusions:** These data provide further insight into the mechanism of action of metformin, potentially identifying novel targets for diabetes treatment, and highlight the need to account for metformin exposure in proteomic studies and where protein biomarkers are used for clinical care where metformin treatment will generate false positive results.

**Highlights:**

- In the most comprehensive proteomic analysis of metformin exposure to date, we showed 97 proteins to be associated with metformin exposure in at least one study.
- 14 proteins were consistently associated with metformin exposure in 2 or more platforms or studies.
- Gene enrichment analysis shows that the strongest protein set is of intestinal origin.
- These data provide further insight into the mechanism of action of metformin, potentially identify novel targets for diabetes treatment and highlight the need to account for metformin exposure in proteomic studies and where protein biomarkers are used for clinical care.

---

Metformin works by several mechanisms known to have a positive impact on inflammation and metabolism; it does this by primarily acting on the liver and the gut(1), however the exact molecular mechanisms remain uncertain. The increasing availability of deep molecular phenotyping in patients treated with metformin, including genomic, transcriptomic, metabolic, proteomic and metagenomic data, offers the opportunity to gain further mechanistic insight into the mechanism of action of metformin in humans. Genome-wide association studies (GWAS) have provided some insight into the mechanism of action of metformin in people with type 2 diabetes(2). In GWAS studies, two genetic variants have been reproducibly associated with glycaemic response to metformin – rs11212617 at a locus including *ATM*(3) and rs8192675, intronic in *SLC2A2*, associated with altered expression of GLUT2(4). Other studies have reported on epigenetic markers(5), the transcriptome(6), the metabolome(7) and the microbiome(8) altering glycaemic response, weight change or intolerance in people with diabetes.

There are limited proteomic studies of metformin exposure, and these have largely been targeted or using small panels. A reproducible robust association has been described between metformin exposure and serum Growth differentiation factor 15 (GDF15) concentrations.

This was first identified in a Luminex panel measuring 237 proteins from the ORIGIN study(9). This has been subsequently replicated with mechanistic rodent studies establishing that metformin associated increase in GDF15 resulted in a reduction in food intake and body weight and that the origin of GDF15 associated with metformin exposure was the intestine(10). More recently, Gummesson et al carried out a more comprehensive proteomic analysis following metformin treatment. This further showed that GDF15 was increased following metformin treatment in addition to identifying other proteins significantly altered by metformin exposure; for example, EpCAM was reduced in those treated with metformin(11). Given the potential for proteomic signatures after metformin exposure to inform on its mechanism of action and identify novel diabetes drug targets, here we extend these analyses, including the study by Gummesson et al. but greatly increasing the number of individuals included to 2,057 and increasing the number of studies to incorporate both cross-sectional and longitudinal studies of metformin exposure, using two commonly used proteomic methods – Olink and SomaLogic.

## Research Design and Methods

### Cohorts

#### GoDARTS

The Genetics of Diabetes Audit and Research Tayside Study (GoDARTS) is a cohort of ∼8,000 individuals with T2D(12). Laboratory measurements were non-fasted. For SomaLogic analysis, samples from 599 patients were selected age >35 years, GAD antibody negative, with blood sampled close to diagnosis (median diabetes duration 1.4 years).

#### DCS

The Hoorn Diabetes Care System (DCS) cohort is a prospective cohort with currently over 14,000 individuals with routine care data. In 2008–2014, additional blood sampling was done in 5500 participants, who provided written informed consent. These samples were used for this study. For SomaLogic analysis, samples from 576 patients were selected age >35 years, GAD antibody negative, with blood sampled close to diagnosis (median diabetes duration 2.6 years)(13).

#### DIRECT

This cohort included 784 patients with recently diagnosed type 2 diabetes. The mean age at inclusion was 62 years with the youngest 35 years at baseline, which should exclude any individuals with MODY. Participants were diagnosed within two years before recruitment, were on lifestyle and/or metformin treatment only, and had glycated haemoglobin (HbA_1c_) < 60.0 mmol/mol (< 7.6%) within previous three months(14).

#### S3WP-T2D

This study was carried out to elucidate the changes in the proteome in the early stages of diabetes and how the proteome is affected by diabetes treatment including metformin(11). 52 previously undiagnosed patients were identified as having type 2 diabetes from a screening program and as a result, were recruited for the study. Patients were excluded if they had a pre-existing disease which would affect their ability to participate, severe hyperglycaemia needing hospital attention or immediate insulin therapy, or a major surgical procedure or trauma within 4 weeks. Included patients were treated for diabetes via first line therapy; weight management and exercise with or without metformin which was decided by a doctor. Protein levels in the blood were measured at baseline, one month and 3 months. Of the 52 participants, 51 completed the 3 month follow up visit and for 3 patients plasma samples were not available for the 1 month visit. This left data for 48 patients to be analysed.

#### IMPOCT

The IMPOCT study was designed to investigate the impact of the OCT1 genotype and OCT1 inhibiting drugs on an individual’s ability to tolerate metformin. For our analysis, only the data from when individuals were treated with metformin and placebo was utilised, and not data from individuals on OCT1 inhibiting drugs. 38 healthy participants without diabetes were recruited for this study. They were on metformin for 4 weeks, titrated to a max dose of 1000mg BD which they took for the final week of the study. Protein levels in the blood were measured at baseline and after the 4 weeks of metformin treatment.

#### RAMP

The RAMP study was designed to investigate the response of individuals with ataxia telangiectasia to metformin and pioglitazone. For our analysis, we only utilised data from control patients (without ataxia telangiectasia) on metformin (not pioglitazone). 12 non-diabetic, healthy controls, who had never been on metformin before were started on the drug. They were treated with metformin for 8 weeks, titrated to a maximum dose of 1000mg BD which they were on for the final 4 weeks of the study. Protein levels in the blood were measured at baseline and after the 8 weeks of metformin treatment.

### Proteomics assays

We used two complementary affinity proteomics approaches to determine the relative levels of circulating proteins in blood samples(15). Each technology is capable to measure thousands of proteins, but a list of 500 proteins have been described to correlate with high confidence between the two platforms. Cross-sectional data from GoDARTS and DCS was analysed using SomaLogic; 1195 proteins were measured and included in the analysis after standardized QC. Olink panels were used as follows:

#### DIRECT

The proteins were measured on five Olink panels: Cardiometabolic, Cardiovascular II, Cardiovascular III, Development, Metabolism. After proteins were removed following quality control, this left 450 proteins to be analysed.

#### S3WP-T2D

The proteins were measured on eleven Olink panels (Cardiometabolic, Cell Regulation, Cardiovascular II, Cardiovascular III, Development, Immune Response, Oncology II, Inflammation, Metabolism, Neurology, and Organ Damage).

#### IMPOCT

The proteins were measured on five Olink panels (Cardiometabolic, Cardiovascular II, Cardiovascular III, Development and Metabolism).

#### RAMP

These proteins were measured on the same five Olink panels as the IMPOCT study (Cardiometabolic, Cardiovascular II, Cardiovascular III, Development and Metabolism).

After proteins were removed following quality control and to ensure each protein was included in all three studies, 372 proteins were analysed in the combined analysis of S3WP-T2D, IMPOCT and RAMP.

### Statistical methods

#### RHAPSODY: GoDARTS and DCS

We undertook a linear regression for the biomarker as dependent variable, with metformin exposure (Y/N), adjusted for age and gender. This was done for both DCS and GoDARTS and then data were combined using random effects meta-analysis. A Bonferroni correction was applied for the 1195 assays included in the analysis.

In both cohorts we then analysed the protein levels of the proteins significantly associated with metformin exposure, in relation to the daily metformin dose used by the participants. Protein levels were used as endpoints in linear regression analyses and metformin dose as predictor with adjustment for gender, age, BMI and HbA1c levels. Persons not using metformin were excluded prior to this analysis. Metformin dose was binned per 500mg to ease the interpretation of the data. i.e. the beta is the change in protein level per extra 500mg tablet of metformin. Similar results were obtained in analyses where metformin dose was included as a continuous variable.

#### DIRECT

A linear mixed model was applied using the lmer function of the R package lme4. In this model, the Olink NPX data was adjusted by information related to the donor (age at sampling, sex) the sampling event (date, centre) as well as technical aspects (assay plate).

### Combined analysis: S3WP-T2D, IMPOCT and RAMP

The longitudinal data from the S3WP-T2D study described by Gummesson et al(11) were combined with the longitudinal data from two Dundee studies; IMPOCT and RAMP. In these studies, Olink panels were used to measure proteins before and after metformin exposure.

Statistical analysis was performed using R Studio version 4.1.2. Proteins were analysed using linear mixed models, with the R package LmerTest. The metformin dose and study name were used as a fixed effect and the study individual was used as a random effect. Proteins were removed so that the all the proteins present in the combined study data were analysed in each of the three studies. This left 372 proteins to be analysed in the combined study data.

The metformin dose was simplified and allocated a 0 if the individual was not on metformin and a 1 if the individual was on metformin treatment, regardless of the dose. P values were adjusted using the Bonferroni method.

### Adjusting for BMI with metformin exposure in S3WP-T2D and RAMP

It has been shown that the proteome can vary in response to weight gain and weight loss(16), and that metformin is associated with weight loss. Consequently, a further linear mixed model analysis was performed in which BMI was included as a covariate, in two of the three longitudinal cohorts where weight was measured before and after metformin initiation.

The longitudinal data from S3WP-T2D and RAMP was combined, and the same 372 proteins were analysed as before using a linear mixed model. Again, R package LmerTest was used for this statistical analysis on R Studio version 4.1.2. Metformin dose, study name and BMI were used as fixed effects, with the study individual as a random effect. P values were also adjusted using the Bonferroni method.

### Tissue specific gene expression and gene set analysis for proteins altered by metformin exposure

Using Genotype Tissue Expression (GTEx) analysis we explored in what tissues the genes encoding the proteins altered by metformin were expressed. We then used the enrichR package in R to evaluate which tissues were enriched for, based upon the change in proteins in response to metformin treatment in the combined analysis of SCAPIS, IMPOCT and RAMP. Proteins were converted to gene symbols and upregulated and downregulated proteins were tested separately. An adjusted P-value smaller than 0.05 was considered significant.

### Causal inference-pQTL

Protein Quantitative Trait Loci (pQTL) analysis was carried out on 14 proteins that were significantly associated with metformin exposure in two or more studies. pQTLs were obtained from Sun et al. (BioRxiv, 2022). pQTLs *in cis* were filtered based on the UniProt ID. pQTLs associated with proteins were compared to available traits in the OpenGWAS database, but eQTLs, cancer-, peptide-, unknown metabolite traits were excluded. pQTLs and identified traits were harmonised using the *harmonise_data* function in the TwoSampleMR package. eQTLs were obtained from GTEx v8. EQTLs were considered significant if the p-value was below GTEx’s P-value threshold.

## Results

### Cross sectional metformin exposure and the SomaLogic platform

We first assessed differences in protein levels measured using the SomaLogic platform in metformin treated and untreated individuals from two cross-sectional studies as part of the IMI-RHAPSODY consortium, where proteomic analysis was undertaken on two populations with type 2 diabetes close to diagnosis. The baseline characteristics of the two included cohorts, GoDARTS and DCS cohort are shown in Supplementary Table 1. In the meta-analysis of these two datasets, 1195 proteins were analysed in 1175 subjects. After Bonferroni correction, levels of 34 proteins were significantly associated with metformin exposure (Supplementary Table 2). The proteins where metformin exposure was associated with the largest difference in levels were REG4 (Beta=0.698, SE=0.084), GDF15 (Beta=0.657, SE 0.098), PYY (Beta=0.662, SE=0.147) and FGF19 (Beta=-0.519, SE=0.115). Given that this analysis was cross-sectional, and associations may not be causally associated with metformin exposure we investigated the effect of metformin dose on protein concentrations, as a dose effect would support a causal relationship between metformin exposure and protein expression. Of the 34 proteins whose concentration was associated with metformin exposure adjusted for gender, age, BMI and HbA1c, a nominally significant (p<0.05) dose effect was seen for most proteins (25/34). After Bonferroni correction (p<0.05/34) increased metformin dose was associated with increased REG4, GDF15, CDH6 and PYY concentrations and decreased FGF19 and OMD concentrations (Supplementary Table 3).

### Cross Sectional metformin exposure and the Olink platform

We then assessed differences in protein levels measured using the Olink platform in individuals from the IMI-DIRECT cohort of patients with type 2 diabetes diagnosed within the prior 2 years. Baseline characteristics of the IMI-DIRECT cohort are shown in Supplementary Table 1. In this study, 450 proteins were analysed in 784 subjects. After Bonferroni correction, 13 proteins were significantly different between metformin users and non-users (Supplementary Table 4). The largest signal was seen for a reduction in EpCAM, followed by an increase in SPINK1, REG4 and GDF15).

### Longitudinal metformin exposure and the Olink platform

We then analysed protein concentrations longitudinally in individuals before metformin treatment and after metformin initiation in 3 clinical trials. Baseline characteristics of included cohorts are shown in Supplementary Table 1. Individual level data from the S3WP-T2D(11), IMPOCT and RAMP studies were combined, and 372 proteins were analysed in 98 subjects using Olink panels before and during metformin treatment. After Bonferroni correction, 68 proteins (18% of measured proteins) were found to be significantly changed with metformin treatment (Supplementary Table 5) and are represented in the volcano plot (Figure 1). The top 8 most significant proteins are labelled in the figure and are as follows from most significant to least significant: REG4, GDF-15, EpCAM, SPINK1, REG1A, LDL receptor, IGFBP-2 and t-PA. We analysed the longitudinal data in males and females separately (Supplementary Table 10) and show a consistent signal by sex for the top 6 most significant proteins. However, there were differences, with 30 proteins including IGFBP-2 and t-PA being significantly changed following metformin in males but not females. Similarly, CDH5, FAM3C, HAOX1 and CCL15 were significantly changed after metformin in females but not males.

**Figure 1.**
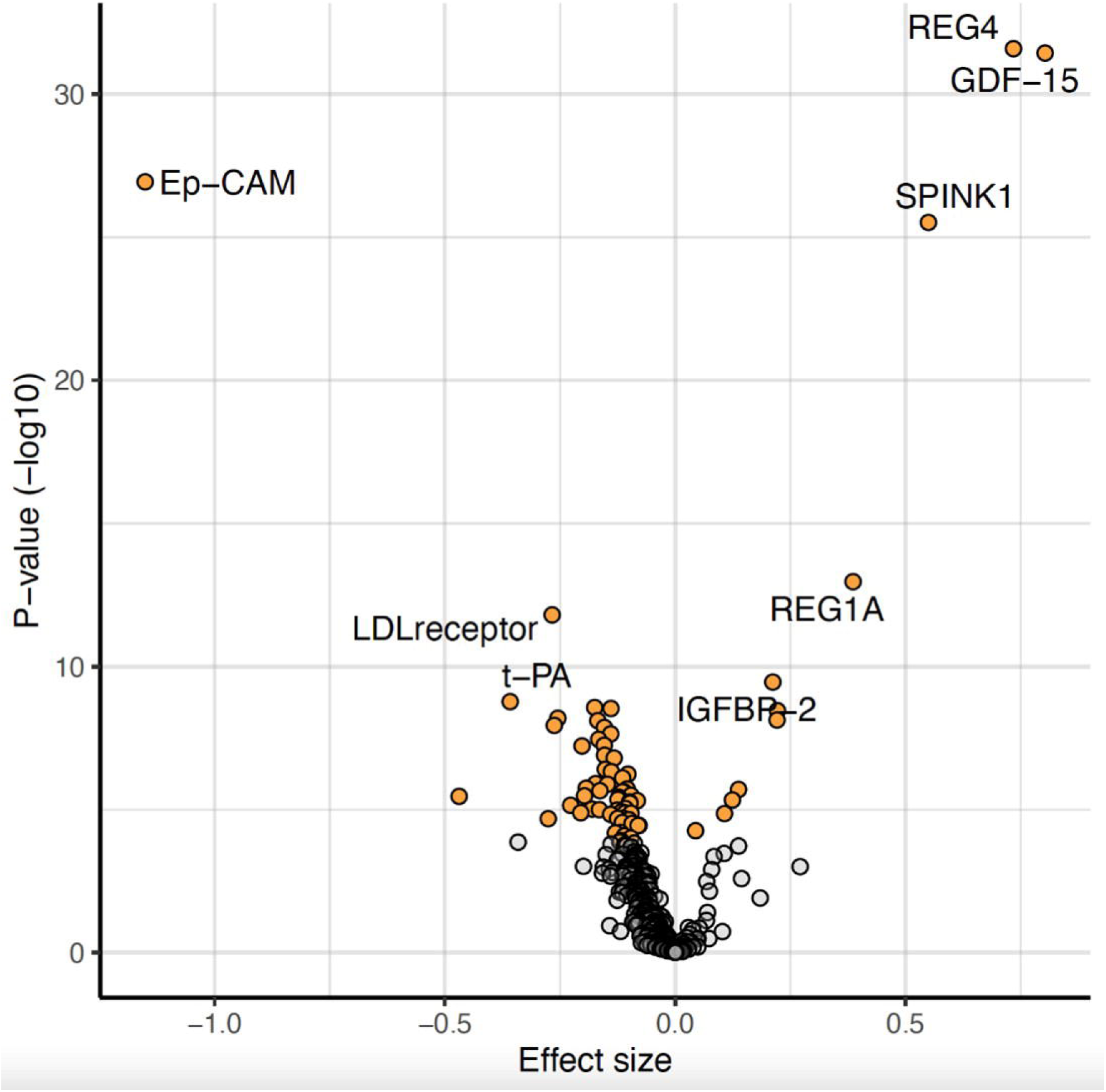
Effect of metformin on plasma protein levels. Volcano plot showing if protein concentrations are significantly increased or decreased following metformin treatment in the longitudinal Olink analysis. Estimate (beta coefficient) is plotted on the x axis and -log10 of the unadjusted p-value (calculated from the linear mixed model) is plotted on the y axis. Proteins with an adjusted p value (Bonferroni method) of less than 0.05 are represented by a yellow dot and all other non-significantly changed proteins are represented by a grey dot. Proteins which have an increased concentration following metformin treatment have a positive effect size whereas proteins which have a decreased concentration following metformin treatment have a negative effect size.

As metformin is associated with weight loss, we then evaluated whether the change in protein concentration with metformin exposure was attenuated by BMI change in two of the longitudinal cohorts where BMI was measured before and after metformin initiation (S3WP-T2D and RAMP). Complete attenuation of the change in protein concentration would suggest that the difference is secondary to, or causal for, BMI change. As a positive control, LEP (Leptin) is significantly reduced by metformin treatment, but adjusting for metformin associated BMI change attenuates the effect by 61%, with loss of significance (p=0.15).

Supplementary Table 9 provides the results for the impact of metformin on protein concentration with and without adjustment for BMI. Adjusting for BMI reduces the number of Bonferroni significant proteins from 31 out of 372 proteins analysed across these two studies to 21 proteins. Other than for leptin, the largest attenuation by adjusting for BMI was seen with FUCA1 (84.2% attenuation); GUSB (29.5% attenuation); SELE (26.3% attenuation), IGFBP2 (21% attenuation) and FCN2 (15% attenuation). Interestingly there was no attenuation (0%) for GDF15, a protein previously reported to potentially mediate the weight change caused by metformin(10).

Given the large number of proteins identified to be altered in our longitudinal analysis we used GTEX(17), HPA(18), and STRING(19) to obtain system-level insights about possible relationships for the 68 proteins identified. The tissue expression (mRNA and protein) from GTEX of the top 68 proteins altered by metformin in the longitudinal analysis is shown in Figure 2. Note that the expression reported here is tissue specific expression and does not relate to metformin exposure. Expression of genes for proteins most changed by metformin exposure were seen predominantly in the pancreas and intestine. In a gene set enrichment analysis (Supplementary Table 7), upregulated proteins were enriched for colon (OR=20.4, Padj = 6.49×10^-5^), based on overlap with REG4, REG1A, GDF15, TFF, SPINK1, CCL15, PIGR and LGALS4. Downregulated proteins were enriched for omentum (OR=5.2, Padj = 2.93×10^-6^) and liver (OR = 4.84, Padj = 7.52×10^-6^). We also explored possible protein interactions using the default functions of the STRING database(19). This revealed that many proteins have known or predicted interactions with each other (Figure 3). Among them, a group of proteins involved in cell adhesion (KEGG pathway, P=2.8×10^-8^) were particularly connected based on experimental evidence.

**Figure 2:**
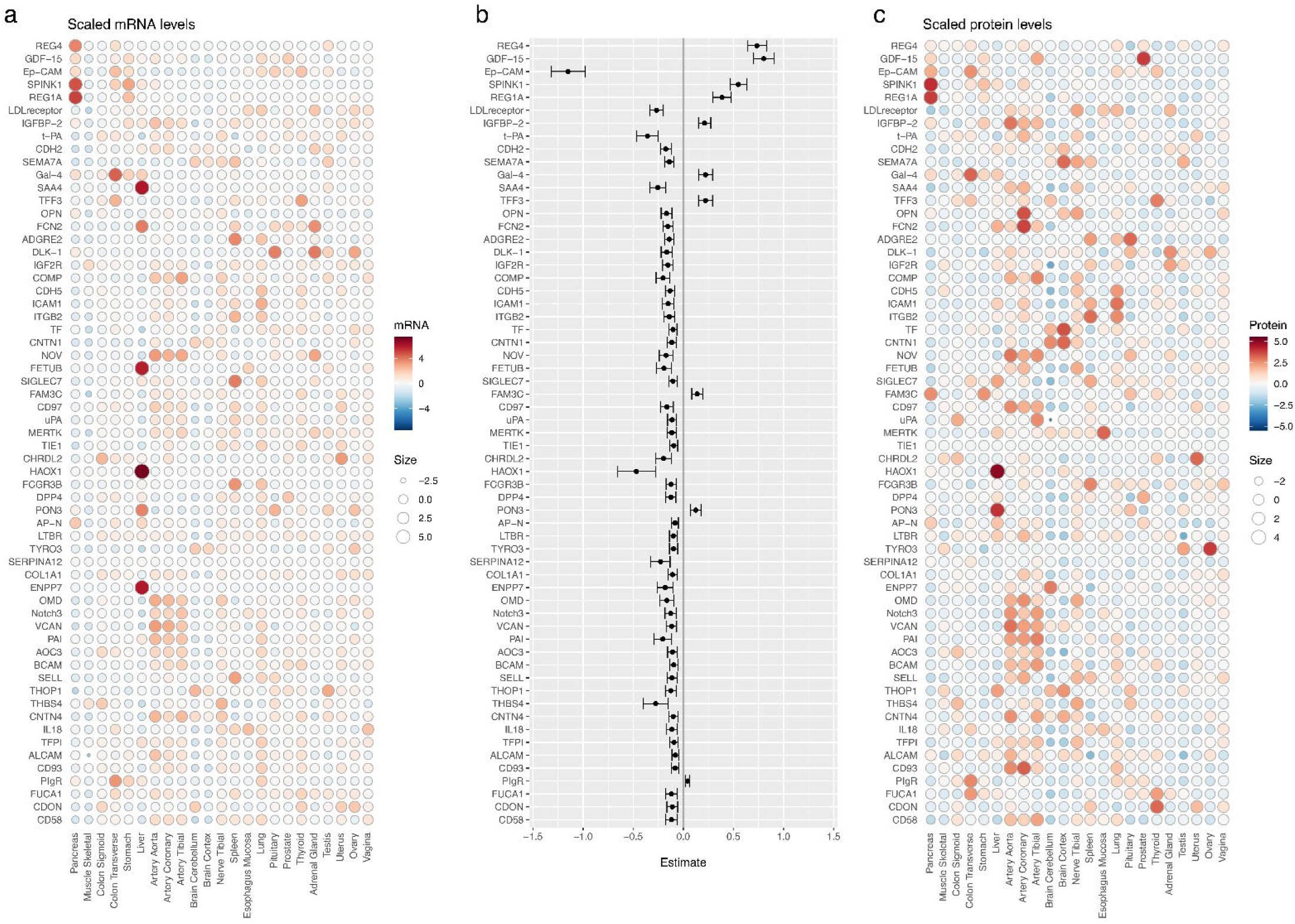
GTEx Analysis Showing the Tissues of Origin of Proteins signficantly altered by metformin exposure. The plot on the left shows the tissues where the mRNA corresponding to the significant proteins are expressed. The plot on the right shows the tissue of origin of the significant proteins. The plot in the middle visualises the significant proteins based upon the longitudinal Olink analysis and whether they are increased or decreased following metformin treatment alongside their tissue of origin. Proteins/gene expression in this figure are ranked in decreasing order of significance. Only 59 of the 68 significant proteins could be included in this analysis due to some proteins missing in the proteomics data. The 9 proteins missing are SEMA7A, GAL-4, LEP, SELE, ADGRG2, CCL15, CD300LG, TIMD4 and PGF.

**Figure 3.**
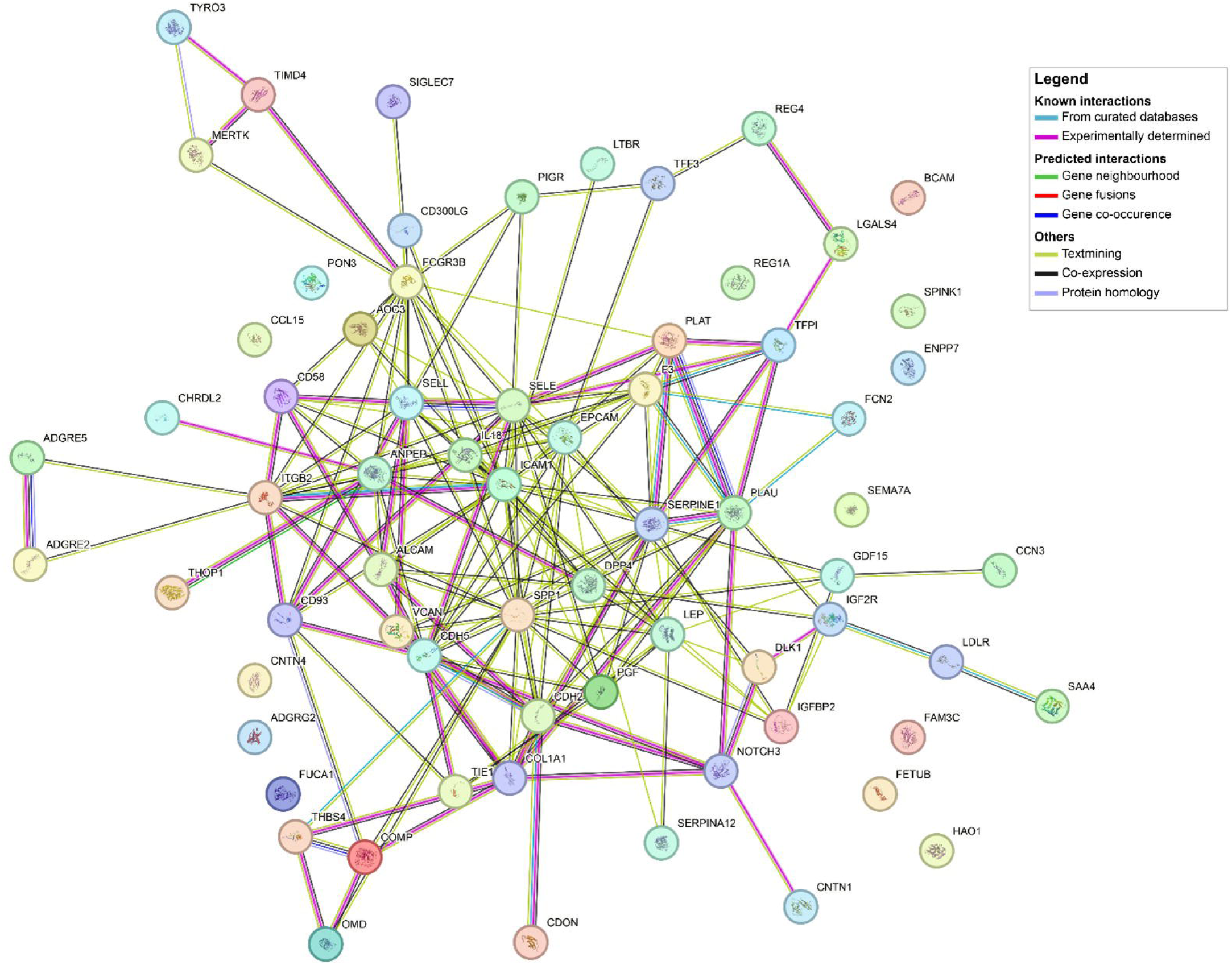
A. Protein relationships for the 68 proteins associated with metformin exposure in the longitudinal Olink study based on StringDB. A large numer of proteins showed experimentally validated interactions shown with with pink lines, were co-expressed (black lines) or were mentioned together (green lines). Experimentally validated interactions are shown in thicker lines (pink, light blue).

### Proteomic signatures of metformin across study and platform

Across the three studies, there were 14 proteins where metformin exposure was associated with protein concentration in at least two studies; the direction and effect sizes for these associations are shown in Figure 4. Four proteins were consistently associated with metformin exposure in the 3 studies (including 2 cross-sectional and 1 longitudinal design) and across the two platforms (Olink and SomaLogic); these were **REG4, GDF15, REG1A and OMD**. 8 additional proteins were consistently associated with metformin exposure across the two studies using Olink; these were **EpCAM, SPINK1, t-PA, Gal-4, TFF3, TF, FAM3C, COL1A1**. There were 2 proteins associated with metformin exposure common between the cross-sectional SomaLogic and longitudinal Olink studies; these were **SELL** and **CD93**. Protein Quantitative Trait Loci (pQTL) analysis was carried out for these 14 proteins. 11 proteins had at least one associated pQTL, while Ep-CAM, SELL and CD93 did not (Supplementary table 8). There were limited informative pQTL: trait associations that could be linked to metformin exposure. A few examples include: where metformin and a pQTL both cause an increase in GDF-15, neutrophil counts are increased, and monocyte counts are decreased; where metformin lowers OMD, the A allele at rs35209758 which is also associated with lower OMD is associated with an increase in Asporin and Hematopoietic progenitor cell antigen CD34; where metformin increases FAM3C, the A allele at rs36198735 that is associated with higher FAM3C is associated with higher heel bone mineral density.

**Figure 4.**
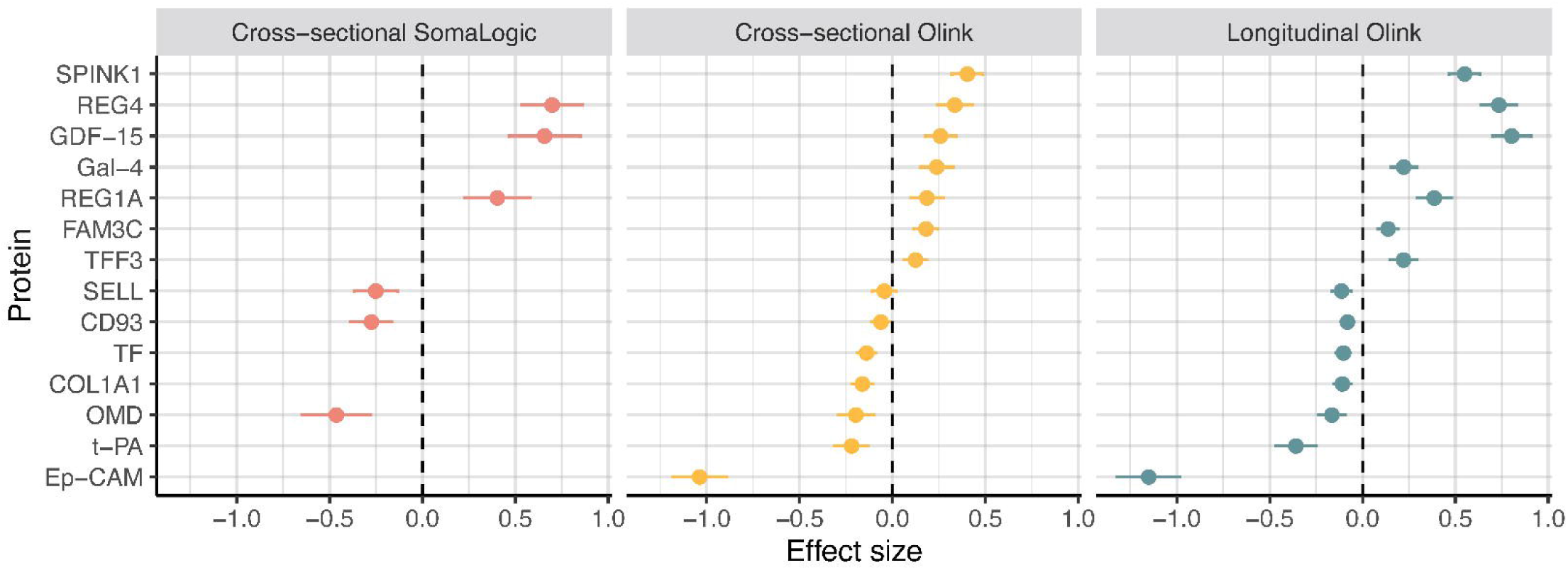
Comparison of effect size across the three studies. (Only proteins that are shared in at least two studies are shown). X-axis, effect size; y-axis, protein.

## Conclusions

We have undertaken the most comprehensive proteomic analysis of metformin exposure in people with and without diabetes to date. Our analysis spans different proteomic approaches and large cross-sectional studies and longitudinal studies with measures before and during metformin treatment. Overall, 97 proteins were associated with metformin exposure in at least one study. The concentration of 4 proteins (REG4, GDF15, REG1A and OMD) were associated with metformin exposure across all platforms and studies, and a further 10 proteins were consistently associated with metformin exposure in two independent studies.

Enrichment analysis shows that the strongest protein-set is of intestinal origin, consistent with the very high concentrations of metformin seen in intestinal epithelial cells. An increase in GDF-15(9) and a decrease in EpCAM(11) after metformin has been previously described and our results have confirmed these findings.

Our data add to the already robust literature that metformin increases serum GDF-15. GDF-15 is a protein that increases in concentration due to cellular stress caused by mitochondrial dysfunction, hypoxia, and exercise(20). It has been previously shown that the intestine (particularly the lower small intestine and colon) was a main site of increased GDF-15 expression following metformin treatment(10). Although GDF-15 is associated with adverse outcomes such as increasing age, cancer and cellular stress, pharmacologically increasing GDF-15 could be beneficial. In wild-type mice treated with a high fat diet, metformin prevented weight gain – an effect not seen in mice lacking GDF15 or lacking the GDF receptor (GFRAL1)(10). These results establish in mice that the weight benefit observed with metformin treatment was mediated by metformin associated increase in GDF15. In the CAMERA trial of 74 non-diabetic participants there was a weak correlation between serum GDF15 concentrations and weight loss in metformin treated individuals(10). However, our data do not support a role for GDF15 in mediating the weight benefits of metformin as, unlike for leptin, we show no attenuation of the GDF15 association with metformin when adjusting for weight change.

Our tissue enrichment analysis identified a set of 8 proteins originating from the intestine as the strongest tissue contributing to the metformin signature: **REG4**, **GDF15**, **REG1A**, IGFBP2, **TFF3**, **SPINK1**, **Gal-4**, PIgR, with all but IGFBP2 and PlgR identified in two or more studies. Two are Regenerating gene (REG) proteins – REG4 and REG1A – which are a part of the calcium-dependent (C-type) lectin superfamily(21). These proteins have been shown to be responsible for triggering cellular proliferation and are associated with some malignancies such as colorectal cancer(22). REG4 can act as a marker for both enteroendocrine cells and Paneth cells in the small intestine(23) and deep crypt secretory cells (the colon equivalent of Paneth cells) in the colon(24), and has been shown to modulate intestinal inflammation and is associated with ulcerative colitis and Crohn’s disease(21).

Trefoil Factor Peptide 3 (TFF3) has a role in colonic epithelial homeostasis and response to gastrointestinal inflammation and mucosal injury(25). Increased TFF3 in mouse hepatocytes has been shown to cause inhibition of genes involved in gluconeogenesis such as PEPCK, G6pc and PGC-1α, reducing hepatic glucose output(26). Moreover, adenoviral overexpression of TFF3 was shown to improve glucose tolerance and insulin sensitivity in diabetic mice(26). In addition, TFF3 has been demonstrated to increase beta cell mass in rat pancreatic islets(27). Serine Protease Inhibitor Kazal Type 1 (SPINK 1) is produced by pancreatic acinar cells and has two main functions; acting as a trypsin inhibitor which acts to protect the pancreas and acting as a cell growth and survival factor which leads to tumour progression(28). Its role in pancreas protection is very important as mutations in the SPINK1 gene are associated with different forms of chronic pancreatitis(29).

The strong association of these intestinal-related proteins with metformin treatment may simply reflect the high exposure of intestinal epithelial cells to metformin and does not necessarily implicate these proteins as mediating any beneficial or potentially harmful effects of metformin. A pQTL association with a trait could help inform on any causal benefit or harm, although a pQTL in a non-metformin exposed state may differ from a pQTL under metformin treatment. For example, metformin increases GDF-15 and a pQTL SNP associated with increased serum GDF15 (in population level data) was associated with increased neutrophil and lower lymphocytes. This is consistent with GDF15 being associated with adverse conditions (age, cancer, cellular stress) but not consistent with the known effect of metformin to lower neutrophil:lymphocyte ratio(30). Thus, although we do find pQTL associated traits, these need to be interpreted with caution. However, whilst we cannot conclude that the intestinal signature for the metformin proteome mediates metformin, it is important to be aware of these strong associations as they could potentially be major confounders in any proteomic analysis where some people may be metformin treated, and clinically where a protein concentration is being used as a clinical biomarker, such as a tumour marker. For example, REG4 is a postulated tumour marker for pancreatic adenocarcinoma(31), gastric and colorectal cancer(32), EpCAM is a well-known tumour marker associated with many cancers including colorectal, ovarian and breast cancers(33) and REG1A has been recently associated with the development of pancreatic cancer(34).

The use of two proteomic platforms in both cross sectional and interventional studies, totalling 2057 participants is a major strength of our study. However, we recognise there are limitations. Firstly, newer proteomic panels include substantially more proteins (e.g. Olink Explore HT measures >5300 proteins, and Somascan measures 7000 proteins). Secondly whilst we combine 3 studies that measure proteins before and after metformin initiation, the number of participants in these longitudinal studies remains small, especially when considering the impact of metformin on BMI change. Finally, whilst we establish a large number of robust signals and for some infer causal association, we don’t establish a causal mechanism for any of the associations. This will require further work with, for example, mouse models as has been demonstrated for the mechanistic contribution of GDF15 to metformin action(10).

In conclusion, we have carried out a comprehensive study on changes to the proteome following metformin treatment. We identified many proteins that are increased or decreased with metformin treatment, with enrichment for proteins originating from the intestine.

Overall, we have shown that the proteomic signature of metformin provides further insight into the mechanism of action, as well as highlighting the potential for false positive signals in human proteomic studies if metformin treatment is not considered as an exposure.

## Data Availability

All data produced in the present study are available upon reasonable request to the authors.

## Acknowledgements

We thank Ragna Häussler, Matilda Dale, the Affinity Proteomics Unit at SciLifeLab in Stockholm for generating the Olink data. This project has received funding from the Innovative Medicines Initiative Joint Undertaking 2, under grant agreement no. 115881 (RHAPSODY) and from the Innovative Medicines Initiative Joint Undertaking under grant agreement no. 115317 (DIRECT), resources of which are composed of financial contribution from the European Union’s Seventh Framework Programme (FP7/2007-2013) and EFPIA companies in kind contribution. There are no relevant conflicts of interest to disclose.

## Author contributions

BC, LM, RS, JMS, AG, LtH, ERP designed the study and wrote the manuscript. BC, KB, LD, JK, RS, LtH analysed the data. MH, RK, PF and all co-authors critically reviewed the final manuscript. ERP is the guarantor of this work and, as such, had full access to all the data in the study and takes responsibility for the integrity of the data and the accuracy of the data analysis.

## Data Availability

All summary results for all 3 studies are provided in the supplementary excel file. The generated individual level proteomic data and linked clinical data in RHAPSODY (DCS, GoDARTS), DIRECT, S3WP-T2D, IMPOCT and RAMP are considered sensitive patient data and cannot be made publicly available in compliance with the European privacy regulations governed by GDPR and according to limitations included in the informed consents signed by the study participants. Data are available by request to the corresponding authors. Requests should include name and contact details of the person requesting the data, which molecular data and clinical variables are requested and the purpose of requesting the data.

## Notes

### Competing Interest Statement

The authors have declared no competing interest.

### Clinical Trial

NCT02586636
NCT02733679

### Funding Statement

This study was funded by the innovative medicines initiative.

### Author Declarations

GoDARTS The study was approved by the East of Scotland Research Ethics Service, reference 09/S1402/44 DCS The study has been approved by the ethics committee of the Amsterdam University Medical Center, location VUmc (approval number 2007/57; NL14692.029.07) DIRECT The UK National Research Ethics Service (NRES) Committee North East Newcastle & North Tyneside gave ethical approval for this work under REC reference 12/NE/0132, Protocol Number 2011DIRECT02 in May 2012. IMPOCT East of Scotland Research Ethics Service gave University of Dundee ethical approval for this work, REC reference: 15/ES/0189 RAMP East of Scotland Research Ethics Service gave University of Dundee ethical approval for this work, REC reference: 16/ES/0056 S3WPT2D The Ethical Review Board in Gothenburg gave ethical approval for this work (Dnr 448-16).

