## Supplementary data for "The Effect of Metformin Treatment on the Circulating Proteome"

**Supplementary figures and tables**

S Figure 1. Study flow chart detailing the 3 separate studies to investigate association of metformin exposure with serum protein concentrations.

S Table 1 – Characteristics of cohort participants.

S Table 2 – Cross-sectional Somalogic analysis: significant proteins associated with metformin treatment.

S Table 3 – Association between metformin dose and protein levels for the 34 proteins associated with metformin treatment in the Cross-sectional Somalogic analysis.

S Table 4 – Cross-sectional Olink analysis: significant proteins associated with metformin treatment.

S Table 5 - Longitudinal Olink analysis of S3WP-T2D, IMPOCT and RAMP: significant proteins associated with metformin treatment.

S Table 6 - Significantly changed proteins following metformin exposure in each of the 3 analyses.

S Table 7 - Gene set enrichment analysis by tissue for proteins significantly up and down regulated by metformin exposure.

S Table 8 - pQTL analysis for top proteins altered by metformin treatment.

S Table 9 - Significantly changed proteins in S3WP-T2D and RAMP following adjustment for BMI.

S Table 10 - Comparison of Longitudinal Olink analysis between males and females.

S Figure 1

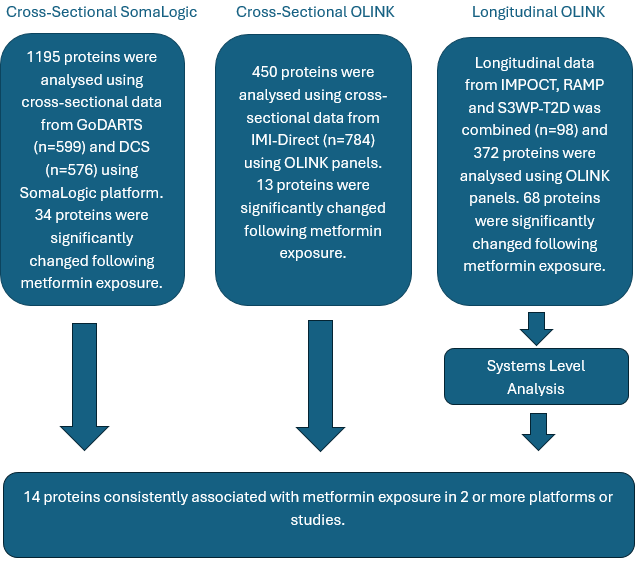

*Flowchart showing the study design which identifies many proteins that are significantly changed following exposure to metformin.*

The two platforms on which proteins were measured are:

**SomaLogic-** This SomaScan assay uses Slow Off-Rate Modified Aptamers (SOMA reagents) and readout on DNA microarrays to determine protien levels as relative fluorescences units (RFU). Information can be found at: <https://somalogic.com/somascan-platform/#:~:text=The%20SomaScan%C2%AE%20Assay%20uses,plasma%2C%20serum%2C%20or%20urine>

**Olink-** Olink’s proximity extension assays (PEA) use two antibodies coupled with complementary oligonucleotides that bind to a common protein target. Followed by qPCR readout, the assays measures 96 proteins per panel. The resulting data is presented in an arbitrary unit called Normalised Protein eXpression (NPX) on a log 2 scale. Information can be found at: <https://www.bioxpedia.com/olink-proteomics/>

S Table 1. Characteristics of cohort participants.

|  | **Cross sectional Somalogic** | | **Cross sectional Olink** | **Longitudinal Olink** | | |
| --- | --- | --- | --- | --- | --- | --- |
| **Clinical Variable** | **GoDarts** | **DCS** | **IMI-DIRECT** | **S3WP-T2D** | **IMPOCT** | **RAMP** |
| Number of Participants, n | 599 | 576 | 784 | 48 | 38 | 12 |
| Type 2 Diabetes Present, Y/N | Y | Y | Y | Y | N | N |
| Number treated with metformin | 266(44.4) | 376 (65.3) | 273 (34.8) | 48 | 38 | 12 |
| Time between metformin initiation and on-treatment samples | Cross sectional | Cross Sectional | Cross Sectional | 1 and 3 months | 4 weeks | 8 weeks |
| Age, years | 61.4(11.0) | 63.2 (10.6) | 61.8 (8.1) | 58.9 | 60.0 | 22.3 |
| Males, n (%) | 354(59.1) | 325 (56.4) | 449 (57.3) | 27 (60.4) | 19 (50) | 6 (50) |
| Body Mass Index, kg/m^2^ | 32.3(6.8) | 30.3 (5.3) | 30.6 (5.1) | 31.1 | 29.9 | 23.1 |

Characteristics of cohort participants. Values are mean unless otherwise stated.

S Table 2. Cross-sectional SomaLogic: significant proteins associated with metformin treatment.

| **SOMA_ID** | **Protein** | **Meta_Beta** | **Meta_SE** | **Meta_zscore** | **Meta_Pvalue** | **Meta_Pvalue_bonf** | **I^2** | **Het_Pvalue** |
| --- | --- | --- | --- | --- | --- | --- | --- | --- |
| SL012561 | REG4 | 0.70 | 0.084 | 8.35 | 6.74E-17 | 8,06E-14 | 0.59 | 0.12 |
| SL003869 | GDF15 | 0.66 | 0.098 | 6.68 | 2.38E-11 | 2,85E-08 | 0.73 | 0.054 |
| SL005230 | UNC5C | 0.35 | 0.054 | 6.59 | 4.41E-11 | 5,27E-08 | 0 | 0.41 |
| SL004661 | ACAN | -0.40 | 0.068 | -5.88 | 4.20E-9 | 5,02E-06 | 0.28 | 0.24 |
| SL003764 | NCAM1 | -0.30 | 0.056 | -5.48 | 4.29E-8 | 5,13E-05 | 0 | 0.54 |
| SL004686 | TNFSF15 | 0.31 | 0.057 | 5.45 | 5.13E-8 | 6,13E-05 | 0 | 0.81 |
| SL011049 | MASP1 | 0.29 | 0.056 | 5.16 | 2.54E-7 | 3,04E-04 | 0 | 0.58 |
| SL003970 | PTH | -0.31 | 0.060 | -5.12 | 2.99E-7 | 3,57E-04 | 0 | 0.57 |
| SL008574 | OMD | -0.46 | 0.093 | -4.96 | 7.01E-7 | 8,38E-04 | 0.64 | 0.095 |
| SL007696 | CD93 | -0.28 | 0.057 | -4.80 | 1.58E-6 | 1,89E-03 | 0 | 0.76 |
| SL010530 | OCIAD1 | 0.27 | 0.057 | 4.78 | 1.75E-6 | 2,09E-03 | 0 | 0.62 |
| SL012769 | FAM3D | 0.29 | 0.060 | 4.73 | 2.24E-6 | 2,68E-03 | 0 | 0.77 |
| SL004636 | FLT3 | 0.26 | 0.057 | 4.60 | 4.20E-6 | 5,01E-03 | 0 | 0.71 |
| SL004804 | CADM3 | 0.26 | 0.057 | 4.54 | 5.70E-6 | 6,81E-03 | 0 | 0.46 |
| SL011069 | PRSS27 | 0.26 | 0.057 | 4.53 | 5.95E-6 | 7,11E-03 | 0 | 0.42 |
| SL004337 | FGF19 | -0.52 | 0.12 | -4.51 | 6.36E-6 | 7,60E-03 | 0.75 | 0.044 |
| SL008909 | LGMN | 0.26 | 0.057 | 4.51 | 6.38E-6 | 7,63E-03 | 0 | 0.63 |
| SL011509 | PYY | 0.66 | 0.147 | 4.51 | 6.41E-6 | 7,66E-03 | 0.85 | 0.010 |
| SL005357 | REG1A | 0.40 | 0.090 | 4.49 | 7.18E-6 | 8,58E-03 | 0.58 | 0.12 |
| SL008810 | NEGR1 | -0.25 | 0.055 | -4.45 | 8.60E-6 | 1,03E-02 | 0 | 0.67 |
| SL004865 | CDH6 | 0.25 | 0.056 | 4.44 | 9.10E-6 | 1,09E-02 | 0 | 0.55 |
| SL016566 | STK17B | 0.25 | 0.057 | 4.39 | 1.14E-5 | 1,36E-02 | 0 | 0.47 |
| SL005491 | OPCML | 0.25 | 0.058 | 4.32 | 1.59E-5 | 1,90E-02 | 0 | 0.44 |
| SL004556 | CD55 | -0.24 | 0.056 | -4.31 | 1.61E-5 | 1,92E-02 | 0 | 0.93 |
| SL004097 | SMAD3 | 0.24 | 0.056 | 4.30 | 1.70E-5 | 2,03E-02 | 0 | 0.46 |
| SL021043 | GDF11 MSTN | -0.24 | 0.057 | -4.27 | 1.96E-5 | 2,34E-02 | 0 | 0.49 |
| SL002823 | SELL | -0.25 | 0.059 | -4.26 | 2.04E-5 | 2,44E-02 | 0 | 0.37 |
| SL010381 | DKKL1 | 0.24 | 0.058 | 4.23 | 2.36E-5 | 2,82E-02 | 0 | 0.42 |
| SL004671 | TNFRSF13C | 0.24 | 0.056 | 4.20 | 2.63E-5 | 3,14E-02 | 0 | 0.37 |
| SL018256 | CHKB | 0.24 | 0.057 | 4.19 | 2.76E-5 | 3,30E-02 | 0 | 0.77 |
| SL007871 | CMPK1 | 0.27 | 0.063 | 4.19 | 2.79E-5 | 3,33E-02 | 0.18 | 0.27 |
| SL000337 | CAPN1 CAPNS1 | -0.25 | 0.059 | -4.18 | 2.87E-5 | 3,42E-02 | 0 | 0.85 |
| SL000324 | C8A C8B C8G | 0.25 | 0.060 | 4.11 | 3.90E-5 | 4,66E-02 | 0 | 0.54 |
| SL003542 | EHMT2 | -0.25 | 0.060 | -4.10 | 4.09E-5 | 4,88E-02 | 0 | 0.55 |

34 Proteins significant after Bonferroni correction from meta-analysis of cross-sectional analysis of DCS and GoDARTS (as part of the RHAPSODY consortium), measured using SomaLogic platform and adjusted for age and gender. Blue rows are significant in longitudinal Olink analysis.

S Table 3. Association between metformin dose and protein levels for the top 34 proteins in the cross sectional meta-analysis of DCS and GoDARTS using the somalogic platform. Data are result of fixed effect meta-analysis.

| **SOMA_ID** | **Protein** | **Meta_Beta** | **Meta_SE** | **Meta_Pvalue** | **I^2** | **Het_Pvalue** |
| --- | --- | --- | --- | --- | --- | --- |
| SL012561 | REG4 | 0.22 | 0.026 | 7.69E-18 | 0 | 9.36E-1 |
| SL003869 | GDF15 | 0.22 | 0.025 | 9.65E-18 | 0 | 7.80E-1 |
| SL004337 | FGF19 | -0.18 | 0.026 | 1.44E-12 | 0 | 9.00E-1 |
| SL008574 | OMD | -0.13 | 0.029 | 5.93E-6 | 0 | 7.36E-1 |
| SL004865 | CDH6 | 0.10 | 0.028 | 2.50E-4 | 0 | 9.14E-1 |
| SL011509 | PYY | 0.099 | 0.030 | 7.97E-4 | 0 | 9.50E-1 |
| SL004636 | FLT3 | 0.087 | 0.028 | 1.80E-3 | 0 | 4.00E-1 |
| SL012769 | FAM3D | 0.097 | 0.032 | 2.26E-3 | 0.81 | 2.08E-2 |
| SL004097 | SMAD3 | 0.083 | 0.028 | 2.66E-3 | 0 | 3.20E-1 |
| SL004661 | ACAN | -0.082 | 0.028 | 3.60E-3 | 0 | 7.35E-1 |
| SL011049 | MASP1 | 0.077 | 0.027 | 4.48E-3 | 0 | 4.69E-1 |
| SL004556 | CD55 | -0.076 | 0.027 | 4.95E-3 | 0 | 3.93E-1 |
| SL007871 | CMPK1 | 0.077 | 0.027 | 5.09E-3 | 0 | 5.64E-1 |
| SL016566 | STK17B | 0.075 | 0.027 | 6.13E-3 | 0 | 4.39E-1 |
| SL005491 | OPCML | 0.073 | 0.028 | 8.74E-3 | 0 | 9.87E-1 |
| SL008909 | LGMN | 0.073 | 0.028 | 8.82E-3 | 0 | 5.96E-1 |
| SL003970 | PTH | -0.076 | 0.030 | 1.11E-2 | 0 | 3.48E-1 |
| SL011069 | PRSS27 | 0.069 | 0.028 | 1.29E-2 | 0 | 8.57E-1 |
| SL010530 | OCIAD1 | 0.069 | 0.029 | 1.62E-2 | 0.18 | 2.69E-1 |
| SL005357 | REG1A | 0.071 | 0.030 | 1.71E-2 | 0 | 4.42E-1 |
| SL002823 | SELL | -0.071 | 0.031 | 2.19E-2 | 0 | 7.65E-1 |
| SL004804 | CADM3 | 0.064 | 0.029 | 2.98E-2 | 0 | 5.85E-1 |
| SL007696 | CD93 | -0.060 | 0.028 | 3.33E-2 | 0 | 8.56E-1 |
| SL000324 | C8A C8B C8G | 0.062 | 0.029 | 3.59E-2 | 0 | 6.38E-1 |
| SL021043 | GDF11 MSTN | -0.058 | 0.029 | 4.74E-2 | 0.40 | 1.96E-1 |
| SL008810 | NEGR1 | -0.048 | 0.028 | 8.09E-2 | 0.22 | 2.56E-1 |
| SL018256 | CHKB | 0.048 | 0.028 | 8.96E-2 | 0 | 8.82E-1 |
| SL005230 | UNC5C | 0.033 | 0.026 | 2.17E-1 | 0 | 9.87E-1 |
| SL004671 | TNFRSF13C | 0.035 | 0.029 | 2.26E-1 | 0 | 8.08E-1 |
| SL003764 | NCAM1 | -0.032 | 0.027 | 2.41E-1 | 0 | 6.15E-1 |
| SL003542 | EHMT2 | -0.024 | 0.029 | 4.05E-1 | 0 | 8.69E-1 |
| SL004367 | DKK1 | -0.022 | 0.030 | 4.77E-1 | 0.82 | 1.99E-2 |
| SL000337 | CAPN1 CAPNS1 | -0.016 | 0.030 | 5.80E-1 | 0.71 | 6.48E-2 |
| SL004686 | TNFSF15 | 0.003 | 0.031 | 9.32E-1 | 0 | 9.40E-1 |

Protein levels were used as endpoints in linear regression analyses and metformin dose as predictor with adjustment for gender, age, BMI and HbA1c levels. Proteins levels log_e_ transformed and Z-scaled prior to the analysis. Persons not using metformin were excluded prior to this analysis. Metformin dose was binned per 500mg to ease the interpretation of the data. i.e. the beta is the in- or decrease in protein level per extra 500mg tablet of metformin. Highly similar results were obtained in analyses where metformin dose was included as a continuous variable. Blue results arsignificant in Longitudinal Olink analysis.

S Table 4 – Cross sectional Olink: significant proteins associated with metformin treatment

| Panel | OlinkID | Protein | Uniprot | ChiSq | Beta | Pvalue | P.adjusted |
| --- | --- | --- | --- | --- | --- | --- | --- |
| CVD3 | OID00610 | Ep-CAM | P16422 | 178.2 | -1.04 | 1.17E-40 | 5.19E-38 |
| DEV | OID01415 | SPINK1 | P00995 | 90.2 | 0.41 | 2.19E-21 | 9.72E-19 |
| MET | OID01184 | REG4 | Q9BYZ8 | 48.1 | 0.34 | 4.14E-12 | 1.84E-09 |
| CVD3 | OID00595 | GDF-15 | Q99988 | 35.6 | 0.26 | 2.47E-09 | 1.10E-06 |
| CVD3 | OID00641 | COL1A1 | P02452 | 32.5 | -0.17 | 1.21E-08 | 5.37E-06 |
| CVD2 | OID00399 | TF | P13726 | 30.0 | -0.14 | 4.37E-08 | 1.94E-05 |
| MET | OID01175 | FAM3C | Q92520 | 29.7 | 0.18 | 5.11E-08 | 2.27E-05 |
| CVD3 | OID00626 | Gal-4 | P56470 | 25.2 | 0.23 | 5.13E-07 | 2.28E-04 |
| CVD3 | OID00635 | t-PA | P00750 | 23.4 | -0.23 | 1.34E-06 | 5.95E-04 |
| CAM | OID01245 | CDH1 | P12830 | 22.7 | 0.14 | 1.93E-06 | 8.57E-04 |
| CAM | OID01231 | REG1A | P05451 | 17.0 | 0.19 | 3.83E-05 | 0.02 |
| DEV | OID01482 | OMD | Q99983 | 15.5 | -0.20 | 8.25E-05 | 0.04 |
| CVD3 | OID00573 | TFF3 | Q07654 | 15.5 | 0.12 | 8.33E-05 | 0.04 |

Cross sectional analysis in IMI-DIRECT using 5 Olink panels (450 proteins). 13 proteins are significantly different between Metformin users and non-users.

S Table 5. Longitudinal Olink analysis of S3WP-T2D, IMPOCT and RAMP

| Protein | Estimate | SE | DF | t value | P value | Adjusted P value |
| --- | --- | --- | --- | --- | --- | --- |
| REG4 | 0.73 | 0.048 | 150.8 | 15.2 | 2.54E-32 | 9.45E-30 |
| GDF-15 | 0.80 | 0.053 | 152.6 | 15.1 | 3.60E-32 | 1.34E-29 |
| Ep-CAM | -1.15 | 0.086 | 155.7 | -13.4 | 1.15E-27 | 4.28E-25 |
| SPINK1 | 0.55 | 0.042 | 150.8 | 12.9 | 3.02E-26 | 1.12E-23 |
| REG1A | 0.39 | 0.047 | 153.4 | 8.17 | 1.09E-13 | 4.05E-11 |
| LDLreceptor | -0.27 | 0.035 | 146.2 | -7.73 | 1.58E-12 | 5.88E-10 |
| IGFBP-2 | 0.21 | 0.032 | 147.5 | 6.73 | 3.52E-10 | 1.31E-7 |
| t-PA | -0.36 | 0.056 | 150.0 | -6.42 | 1.70E-9 | 6.32E-7 |
| CDH2 | -0.17 | 0.028 | 145.8 | -6.34 | 2.72E-9 | 1.01E-6 |
| SEMA7A | -0.14 | 0.022 | 146.2 | -6.32 | 2.95E-9 | 1.10E-6 |
| Gal-4 | 0.22 | 0.035 | 148.1 | 6.28 | 3.52E-9 | 1.31E-6 |
| SAA4 | -0.25 | 0.041 | 150.9 | -6.16 | 6.46E-9 | 2.40E-6 |
| TFF3 | 0.22 | 0.036 | 146.9 | 6.14 | 7.44E-9 | 2.77E-6 |
| OPN | -0.17 | 0.027 | 147.8 | -6.12 | 7.86E-9 | 2.92E-6 |
| LEP | -0.26 | 0.043 | 145.8 | -6.05 | 1.15E-8 | 4.28E-6 |
| FCN2 | -0.15 | 0.026 | 146.7 | -6.02 | 1.36E-8 | 5.06E-6 |
| ADGRE2 | -0.14 | 0.024 | 146.9 | -5.91 | 2.28E-8 | 8.48E-6 |
| DLK-1 | -0.17 | 0.029 | 146.8 | -5.82 | 3.53E-8 | 1.31E-5 |
| IGF2R | -0.15 | 0.027 | 146.5 | -5.72 | 5.70E-8 | 2.12E-5 |
| COMP | -0.20 | 0.035 | 151.3 | -5.70 | 6.02E-8 | 2.24E-5 |
| SELE | -0.15 | 0.028 | 146.8 | -5.55 | 1.27E-7 | 4.72E-5 |
| CDH5 | -0.13 | 0.024 | 148.7 | -5.50 | 1.59E-7 | 5.92E-5 |
| ICAM1 | -0.15 | 0.029 | 152.0 | -5.30 | 3.93E-7 | 0.00015 |
| ITGB2 | -0.14 | 0.026 | 146.3 | -5.27 | 4.76E-7 | 0.00018 |
| TF | -0.10 | 0.020 | 145.6 | -5.23 | 5.82E-7 | 0.00022 |
| CNTN1 | -0.11 | 0.022 | 148.0 | -5.16 | 7.91E-7 | 0.00029 |
| NOV | -0.17 | 0.034 | 150.6 | -5.05 | 1.24E-6 | 0.00046 |
| ADGRG2 | -0.15 | 0.029 | 155.2 | -5.03 | 1.31E-6 | 0.00049 |
| FETUB | -0.19 | 0.039 | 151.1 | -4.97 | 1.81E-6 | 0.00067 |
| SIGLEC7 | -0.10 | 0.021 | 147.0 | -4.96 | 1.92E-6 | 0.00071 |
| FAM3C | 0.14 | 0.028 | 151.3 | 4.95 | 1.99E-6 | 0.00074 |
| CD97 | -0.16 | 0.033 | 149.6 | -4.92 | 2.21E-6 | 0.00082 |
| uPA | -0.11 | 0.023 | 148.6 | -4.91 | 2.39E-6 | 0.00089 |
| MERTK | -0.11 | 0.023 | 146.4 | -4.91 | 2.41E-6 | 0.00090 |
| TIE1 | -0.10 | 0.020 | 155.1 | -4.83 | 3.21E-6 | 0.0012 |
| CHRDL2 | -0.20 | 0.041 | 150.0 | -4.83 | 3.34E-6 | 0.0012 |
| HAOX1 | -0.47 | 0.097 | 148.1 | -4.82 | 3.47E-6 | 0.0013 |
| FCGR3B | -0.12 | 0.026 | 147.2 | -4.81 | 3.76E-6 | 0.0014 |
| DPP4 | -0.12 | 0.026 | 150.1 | -4.76 | 4.45E-6 | 0.0017 |
| PON3 | 0.12 | 0.026 | 147.0 | 4.76 | 4.67E-6 | 0.0017 |
| AP-N | -0.08 | 0.017 | 147.7 | -4.74 | 4.91E-6 | 0.0018 |
| LTBR | -0.10 | 0.021 | 149.8 | -4.72 | 5.48E-6 | 0.0020 |
| TYRO3 | -0.10 | 0.021 | 148.6 | -4.69 | 6.03E-6 | 0.0022 |
| SERPINA12 | -0.23 | 0.049 | 146.3 | -4.66 | 7.14E-6 | 0.0027 |
| COL1A1 | -0.11 | 0.024 | 147.8 | -4.59 | 9.25E-6 | 0.0034 |
| ENPP7 | -0.18 | 0.039 | 145.4 | -4.59 | 9.64E-6 | 0.0036 |
| OMD | -0.16 | 0.036 | 150.2 | -4.57 | 1.01E-5 | 0.0038 |
| Notch3 | -0.13 | 0.028 | 150.6 | -4.56 | 1.06E-5 | 0.0039 |
| VCAN | -0.12 | 0.026 | 152.8 | -4.53 | 1.20E-5 | 0.0045 |
| PAI | -0.21 | 0.046 | 155.5 | -4.50 | 1.30E-5 | 0.0048 |
| AOC3 | -0.11 | 0.024 | 150.9 | -4.50 | 1.33E-5 | 0.0049 |
| BCAM | -0.10 | 0.021 | 145.5 | -4.51 | 1.35E-5 | 0.0050 |
| CCL15 | 0.11 | 0.024 | 147.1 | 4.50 | 1.39E-5 | 0.0052 |
| CD300LG | -0.14 | 0.031 | 148.7 | -4.48 | 1.47E-5 | 0.0055 |
| SELL | -0.11 | 0.026 | 149.1 | -4.40 | 2.02E-5 | 0.0075 |
| THOP1 | -0.13 | 0.028 | 150.4 | -4.40 | 2.05E-5 | 0.0076 |
| THBS4 | -0.28 | 0.063 | 150.7 | -4.39 | 2.10E-5 | 0.0078 |
| CNTN4 | -0.11 | 0.023 | 147.0 | -4.38 | 2.27E-5 | 0.0084 |
| IL18 | -0.11 | 0.027 | 146.9 | -4.32 | 2.85E-5 | 0.011 |
| TFPI | -0.09 | 0.022 | 146.7 | -4.30 | 3.08E-5 | 0.011 |
| ALCAM | -0.08 | 0.018 | 147.1 | -4.26 | 3.60E-5 | 0.013 |
| CD93 | -0.08 | 0.019 | 148.0 | -4.26 | 3.62E-5 | 0.013 |
| PIgR | 0.04 | 0.011 | 149.4 | 4.15 | 5.51E-5 | 0.020 |
| FUCA1 | -0.12 | 0.029 | 145.7 | -4.12 | 6.36E-5 | 0.024 |
| TIMD4 | -0.13 | 0.032 | 149.7 | -4.11 | 6.60E-5 | 0.025 |
| CDON | -0.11 | 0.027 | 154.1 | -4.042 | 8.34E-5 | 0.031 |
| PGF | -0.10 | 0.024 | 147.5 | -4.00 | 0.00010 | 0.037 |
| CD58 | -0.12 | 0.030 | 153.4 | -3.94 | 0.00012 | 0.046 |

In a combined longitudinal analysis across 3 datasets (S3WP-T2D, IMPOCT, RAMP) 68 of 372 proteins analysed with Olink panel were significant after Bonferroni correction. Those highlighted in red were significant in the cross-sectional Olink analysis (S Table 3).

S Table 6. Significantly changed proteins following metformin exposure in each of the 3 analyses

| **Protein** | **Confidence Tier** | **Longitudinal OLINK BETA** | **Longitudinal OLINK P VALUE** | **Cross-sectional OLINK BETA** | **Cross-sectional OLINK P VALUE** | **Cross-sectional SomaLogic BETA** | **Cross-sectional SomaLogic P VALUE** |
| --- | --- | --- | --- | --- | --- | --- | --- |
| **REG4** | **1** | **0.73** | **2.54E-32** | **0.34** | **4.14E-12** | **0.70** | **6.74E-17** |
| **GDF-15** | **1** | **0.80** | **3.60E-32** | **0.26** | **2.47E-9** | **0.66** | **2.38E-11** |
| **EP-CAM** | **3** | **-1.15** | **1.15E-27** | **-1.04** | **1.17E-40** |  |  |
| **SPINK1** | **2** | **0.55** | **3.02E-26** | **0.41** | **2.19E-21** |  |  |
| **REG1A** | **1** | **0.39** | **1.09E-13** | **0.19** | **3.83E-5** | **0.40** | **7.18E-6** |
| **LDLRECEPTOR** | **3** | **-0.27** | **1.58E-12** | 0.047 | 0.33 | -0.048 | 7.28E-1 |
| **IGFBP-2** | **1** | **0.21** | **3.52E-10** | -0.044 | 0.37 |  |  |
| **T-PA** | **1** | **-0.36** | **1.70E-9** | **-0.23** | **1.34E-6** |  |  |
| **CDH2** | **3** | **-0.17** | **2.72E-9** | 0.083 | 0.051 | -0.088 | 1.50E-1 |
| **SEMA7A** | **2** | **-0.14** | **2.95E-9** | -0.030 | 0.32 |  |  |
| **GAL-4** | **2** | **0.22** | **3.52E-9** | **0.23** | **5.13E-7** | 0.072 | 2.37E-1 |
| **SAA4** | **2** | **-0.25** | **6.46E-9** | -0.12 | 0.018 |  |  |
| **TFF3** | **1** | **0.22** | **7.44E-9** | **0.12** | **8.33E-5** | 0.19 | 5.74E-4 |
| **OPN** | **1** | **-0.17** | **7.86E-9** | -0.088 | 0.044 |  |  |
| **LEP** | **1** | **-0.26** | **1.15E-8** | 0.012 | 0.85 | 0.042 | 6.60E-1 |
| **FCN2** | **1** | **-0.15** | **1.36E-8** | -0.027 | 0.55 | 0.16 | 5.25E-3 |
| **ADGRE2** | **1** | **-0.14** | **2.28E-8** | 0.0025 | 0.94 |  |  |
| **DLK-1** | **1** | **-0.17** | **3.53E-8** | -0.083 | 0.11 |  |  |
| **IGF2R** | **1** | **-0.15** | **5.70E-8** | 0.026 | 0.36 | -0.13 | 2.84E-2 |
| **COMP** | **1** | **-0.20** | **6.02E-8** | -0.14 | 0.00066 |  |  |
| **SELE** | **1** | **-0.15** | **1.27E-7** | 0.026 | 0.59 | 0.12 | 1.05E-1 |
| **CDH5** | **1** | **-0.13** | **1.59E-7** | -0.041 | 0.019 | -0.29 | 2.54E-4 |
| **ICAM1** | **2** | **-0.15** | **3.93E-7** | 0.0054 | 0.87 | 0.066 | 6.50E-1 |
| **ITGB2** | **3** | **-0.14** | **4.76E-7** | -0.10 | 0.00039 |  |  |
| **TF** | **2** | **-0.10** | **5.82E-7** | **-0.14** | **4.37E-8** | -0.026 | 6.69E-1 |
| **CNTN1** | **1** | **-0.11** | **7.91E-7** | -0.063 | 0.017 | -0.20 | 8.07E-4 |
| **NOV** | **2** | **-0.17** | **1.24E-6** | -0.074 | 0.024 | 0.080 | 1.97E-1 |
| **ADGRG2** | **3** | **-0.15** | **1.31E-6** | -0.073 | 0.0017 |  |  |
| **FETUB** | **1** | **-0.19** | **1.81E-6** | -0.011 | 0.75 | -0.12 | 4.05E-2 |
| **SIGLEC7** | **2** | **-0.10** | **1.92E-6** | 0.060 | 0.029 | -0.015 | 8.05E-1 |
| **FAM3C** | **3** | **0.14** | **1.99E-6** | **0.18** | **5.11E-8** |  |  |
| **CD97** | **1** | **-0.16** | **2.21E-6** | 0.013 | 0.71 | 0.098 | 1.07E-1 |
| **UPA** | **2** | **-0.11** | **2.39E-6** | -0.048 | 0.13 |  |  |
| **MERTK** | **3** | **-0.11** | **2.41E-6** | -0.067 | 0.029 |  |  |
| **TIE1** | **2** | **-0.10** | **3.21E-6** | -0.025 | 0.20 | -0.13 | 3.16E-2 |
| **CHRDL2** | **1** | **-0.20** | **3.34E-6** | -0.076 | 0.13 |  |  |
| **HAOX1** | **3** | **-0.47** | **3.47E-6** | 0.24 | 0.030 |  |  |
| **FCGR3B** | **2** | **-0.12** | **3.76E-6** | -0.057 | 0.19 | -0.035 | 5.59E-1 |
| **DPP4** | **2** | **-0.12** | **4.45E-6** | -0.042 | 0.16 |  |  |
| **PON3** | **3** | **0.12** | **4.67E-6** | -0.080 | 0.097 |  |  |
| **AP-N** | **3** | **-0.08** | **4.91E-6** | -0.052 | 0.059 |  |  |
| **LTBR** | **3** | **-0.10** | **5.48E-6** | -0.034 | 0.21 | 0.11 | 7.37E-2 |
| **TYRO3** | **2** | **-0.10** | **6.03E-6** | -0.0089 | 0.67 | -0.077 | 2.09E-1 |
| **SERPINA12** | **1** | **-0.23** | **7.14E-6** | -0.14 | 0.08 |  |  |
| **COL1A1** | **1** | **-0.11** | **9.25E-6** | **-0.17** | **1.21E-8** |  |  |
| **ENPP7** | **1** | **-0.18** | **9.64E-6** | 0.13 | 0.10 | 0.11 | 6.84E-2 |
| **OMD** | **1** | **-0.16** | **1.01E-5** | **-0.20** | **8.25E-5** | **-0.46** | **7.01E-7** |
| **NOTCH3** | **1** | **-0.13** | **1.06E-5** | -0.11 | 0.00058 | -0.22 | 2.37E-4 |
| **VCAN** | **3** | **-0.12** | **1.20E-5** | -0.053 | 0.047 |  |  |
| **PAI** | **1** | **-0.21** | **1.30E-5** | 0.070 | 0.25 |  |  |
| **AOC3** | **3** | **-0.11** | **1.33E-5** | -0.011 | 0.71 |  |  |
| **BCAM** | **2** | **-0.10** | **1.35E-5** | 0.0095 | 0.71 | -0.13 | 3.17E-1 |
| **CCL15** | **1** | **0.11** | **1.39E-5** | 0.027 | 0.50 | 0.15 | 1.05E-2 |
| **CD300LG** | **3** | **-0.14** | **1.47E-5** | -0.044 | 0.20 |  |  |
| **SELL** | **1** | **-0.11** | **2.02E-5** | -0.044 | 0.19 | **-0.25** | **2.04E-5** |
| **THOP1** | **3** | **-0.13** | **2.05E-5** | 0.039 | 0.37 |  |  |
| **THBS4** | **1** | **-0.28** | **2.10E-5** | -0.17 | 0.00049 | -0.29 | 2.15E-2 |
| **CNTN4** | **2** | **-0.11** | **2.27E-5** | -0.016 | 0.57 | -0.099 | 9.76E-2 |
| **IL18** | **3** | **-0.11** | **2.85E-5** | 0.055 | 0.18 |  |  |
| **TFPI** | **2** | **-0.09** | **3.08E-5** | -0.056 | 0.056 | 0.066 | 2.69E-1 |
| **ALCAM** | **2** | **-0.08** | **3.60E-5** | -0.025 | 0.30 | -0.10 | 8.02E-2 |
| **CD93** | **1** | **-0.08** | **3.62E-5** | -0.066 | 0.013 | **-0.28** | **1.58E-6** |
| **PIGR** | **2** | **0.04** | **5.51E-5** | 0.017 | 0.15 | -0.097 | 1.66E-1 |
| **FUCA1** | **3** | **-0.12** | **6.36E-5** | 0.11 | 0.056 |  |  |
| **TIMD4** | **1** | **-0.13** | **6.60E-5** | -0.11 | 0.0068 |  |  |
| **CDON** | **1** | **-0.11** | **8.34E-5** | -0.019 | 0.43 | -0.065 | 2.73E-1 |
| **PGF** | **2** | **-0.10** | **0.0001** | -0.025 | 0.29 | -0.017 | 7.75E-1 |
| **CD58** | **2** | **-0.12** | **0.00012** | -0.0088 | 0.72 |  |  |
| **CDH1** | **3** | 0.14 | 0.00019 | **0.14** | **1.93E-6** | 0.25 | 5.18E-3 |
| **UNC5C** | **3** |  |  |  |  | **0.35** | **4.41E-11** |
| **ACAN** | **2** | -0.078 | 0.00088 | -0.049 | 0.063 | **-0.40** | **4.20E-9** |
| **NCAM1** | **2** | -0.90 | 0.0055 | -0.077 | 0.0091 | **-0.30** | **4.29E-8** |
| **TNFSF15** | **3** |  |  |  |  | **0.31** | **5.13E-8** |
| **MASP1** | **2** |  |  |  |  | **0.29** | **2.54E-7** |
| **PTH** | **3** |  |  |  |  | **-0.31** | **2.99E-7** |
| **OCIAD1** | **3** |  |  |  |  | **0.27** | **1.75E-6** |
| **FAM3D** | **1** |  |  |  |  | **0.29** | **2.24E-6** |
| **FLT3** | **3** |  |  |  |  | **0.26** | **4.20E-6** |
| **CADM3** | **3** |  |  |  |  | **0.26** | **5.70E-6** |
| **PRSS27** | **1** | -0.0083 | 0.70 | 0.0090 | 0.81 | **0.26** | **5.95E-6** |
| **FGF19** | **3** |  |  |  |  | **-0.52** | **6.36E-6** |
| **LGMN** | **1** | -0.035 | 0.19 | 0.047 | 0.15 | **0.26** | **6.38E-6** |
| **PYY** | **1** |  |  |  |  | **0.66** | **6.41E-6** |
| **NEGR1** | **3** |  |  |  |  | **-0.24** | **8.60E-6** |
| **CDH6** | **3** |  |  |  |  | **0.25** | **9.10E-6** |
| **STK17B** | **3** |  |  |  |  | **0.25** | **1.14E-5** |
| **OPCML** | **3** |  |  |  |  | **0.25** | **1.59E-5** |
| **CD55** | **2** |  |  |  |  | **-0.24** | **1.61E-5** |
| **SMAD3** | **3** |  |  |  |  | **0.24** | **1.70E-5** |
| **GDF11MSTN** | **3** |  |  |  |  | **-0.24** | **1.96E-5** |
| **DKKL1** | **2** |  |  |  |  | **0.24** | **2.36E-5** |
| **TNFRSF13C** | **3** |  |  |  |  | **0.24** | **2.63E-5** |
| **CHKB** | **3** |  |  |  |  | **0.24** | **2.76E-5** |
| **CMPK1** | **3** |  |  |  |  | **0.27** | **2.79E-5** |
| **CAPN1CAPNS1** | **3** |  |  |  |  | **-0.25** | **2.87E-5** |
| **C8AC8BC8G** | **3** |  |  |  |  | **0.25** | **3.90E-5** |
| **EHMT2** | **3** |  |  |  |  | **-0.25** | **4.09E-5** |

Supplementary Table 6.

Table showing proteins that were significantly changed after metformin in at least one study. The beta and p value of each protein is shown if the protein is included in that study and highlighted in bold if significant for that particular study. Proteins highlighted in red were significantly associated with metformin exposure in all three studies. Proteins highlighted in blue are significant in 2 studies. Confidence tiers are shown based upon the presence of a cis-pQTL across the OLINK and Somalogic platforms as described in this study (https://doi.org/10.1038/s41586-023-06563-x); where tier 1 had cis-PQTL on two platforms with strong correlation; tier 2 had a cis-pQTL on one platform only or on two platforms but with weak correlation; tier 3 did not have a cis-pQTL on either the OLINK or somalogic platform.

S Table 7. Gene set enrichment analysis by tissue for proteins significantly up and down regulated by metformin exposure.

| **Term** | **Overlap** | **P. Value** | **Adjusted.P.value** | **Odds.Ratio** | **Direction** |
| --- | --- | --- | --- | --- | --- |
| **OMENTUM** | 23/2316 | 2.71E-8 | 2.93E-6 | 5.21 | Downregulated |
| **LIVER (BULK TISSUE)** | 22/2316 | 1.39E-7 | 7.53E-6 | 4.84 | Downregulated |
| **COLON (BULK TISSUE)** | 8/2316 | 3.82E-6 | 6.49E-5 | 20.4 | Upregulated |
| **GASTRIC EPITHELIAL CELL** | 8/2316 | 3.82E-6 | 6.49E-5 | 20.4 | Upregulated |
| **GASTRIC TISSUE (BULK)** | 8/2316 | 3.82E-6 | 6.49E-5 | 20.4 | Upregulated |
| **SMALL INTESTINE (BULK TISSUE)** | 8/2316 | 3.82E-6 | 6.49E-5 | 20.4 | Upregulated |
| **VALVE** | 19/2316 | 1.23E-5 | 0.00044 | 3.84 | Downregulated |
| **ILEUM (BULK)** | 7/2316 | 5.98E-5 | 0.00068 | 13.4 | Upregulated |
| **LIVER (BULK TISSUE)** | 7/2316 | 5.98E-5 | 0.00068 | 13.4 | Upregulated |
| **LUNG (BULK TISSUE)** | 18/2316 | 4.71E-5 | 0.0013 | 3.54 | Downregulated |
| **PLACENTA (BULK)** | 17/2316 | 0.00017 | 0.0030 | 3.26 | Downregulated |
| **RENAL CORTEX** | 17/2316 | 0.00017 | 0.0030 | 3.26 | Downregulated |
| **COLONIC MUCOSA** | 6/2316 | 0.00066 | 0.0056 | 9.18 | Upregulated |
| **HEPATOCYTE** | 6/2316 | 0.00066 | 0.0056 | 9.18 | Upregulated |
| **ADIPOSE (BULK TISSUE)** | 16/2316 | 0.00055 | 0.0059 | 2.99 | Downregulated |
| **BREAST (BULK TISSUE)** | 16/2316 | 0.00055 | 0.0059 | 2.99 | Downregulated |

Protein identifiers were converted to gene symbols. Enrichment was performed using the enrichR package in R. Upregulated proteins and downregulated proteins were tested separately. An adjusted P-value smaller than 0.05 was considered significant. Upregulated proteins were enriched for genes related to the intestine and stomach (OR=20.4, Padj = 6.49x10^-6^) which included eight proteins (REG4, GDF15, REG1A, IGFBP2, TFF3, SPINK1, Gal-4, PIgR). Downregulated proteins were enriched for omentum (OR=5.2, Padj = 2.93x10^-6^), liver (OR = 4.84, Padj = 7.52x10^-6^).

S Table 8. pQTL analysis of proteins consistently associated with metformin exposure

| **rsID** | **βMetf** | **REF** | **ALT** | Protein | β | SE | Log10P | Trait | Beta | P | EA |
| --- | --- | --- | --- | --- | --- | --- | --- | --- | --- | --- | --- |
| rs79795228 | 0.73 | C | A | REG4 | 0.63 | 0.027 | 116.59 | None |  |  |  |
| rs1054221 | 0.80 | T | C | GDF15 | 0.39 | 0.009 | 425.56 | Monocyte percentage | -0.025 | 7.97e-15 | C |
| rs1054221 | 0.80 | T | C | GDF15 | 0.39 | 0.009 | 425.56 | neutrophil cell count | 0.02 | 2.67e-13 | C |
| rs7725017 | 0.55 | C | A | SPINK1 | -0.40 | 0.008 | 628.53 | None |  |  |  |
| rs11126696 | 0.39 | A | G | REG1A | 0.25 | 0.007 | 249.09 | None |  |  |  |
| rs2020921 | -0.36 | G | A | t-PA | -0.28 | 0.022 | 35.36 | None |  |  |  |
| rs140695578 | 0.22 | A | T | Gal-4 | 0.27 | 0.019 | 46.00 | Mean platelet (thrombocyte) volume | -0.03 | 3.21e-7 | T |
| rs118095917 | 0.22 | C | T | TFF3 | -1.25 | 0.045 | 168.75 | Enteropathic arthropathies | 4.39 | 2.26e-4 | T |
| rs6666213 | -0.10 | A | G | TF | 0.18 | 0.007 | 148.62 | Prothrombin time | -0.03 | 1.36e-7 | G |
| rs36198735 | 0.14 | A | AAAC | FAM3C | -0.11 | 0.008 | 41.98 | Heel bone mineral density | -0.04 | 8.20e-34 | AAAC |
| rs147266928 | -0.11 | T | C | COL1A1 | 1.39 | 0.108 | 37.03 | None |  |  |  |
| rs35209758 | -0.16 | A | AT | OMD | 0.08 | 0.008 | 22.95 | Asporin | 0.221 | 3.02e-16 | A |
| rs35209758 | -0.16 | A | AT | OMD | 0.08 | 0.008 | 22.95 | Hematopoietic progenitor cell antigen CD34 | 0.204 | 6.03e-14 | A |

pQTLs were identified for 11 of the 14 proteins consistently associated with metformin exposure. For each pQTL we looked up the P-values and betas of associated traits based on data in the IEU Open GWAS project (https://gwas.mrcieu.ac.uk/) . Thus the table reports 1) how metformin exposure alters the levels of the protein (βmetf, positive if metformin increases the protein), 2) the pQTL SNP association with the protein (β is positive if the alt allele is associated with an increase in the protein) and 3) the association of the pQTL with any associated trait, where Beta represents the association of the effect allele (EA) with the level of the trait.

Supplementary Table 9. Attenuation of metformin association by adjusting for weight change

|  | **Unadjusted for weight change** | | | **Adjusted for weight change** | | |  |
| --- | --- | --- | --- | --- | --- | --- | --- |
| Protein | Estimate | P value | Adj P value | **Estimate** | **P value** | **Adj P Value** | Percentage Attenuation |
| Ep-CAM | -1.25 | 3.41e-22 | 1.27e-19 | **-1.25** | **3.25e-22** | **1.21e-19** | 0.12% |
| GDF-15 | 0.69 | 5.39e-18 | 2.00e-15 | **0.69** | **4.90e-18** | **1.82e-15** | 0.26% |
| REG4 | 0.64 | 1.97e-20 | 7.34e-18 | **0.62** | **6.00e-20** | **2.23e-17** | -2.48% |
| SPINK1 | 0.44 | 4.46e-13 | 1.66e-10 | **0.43** | **1.41e-12** | **5.25e-10** | -2.85% |
| REG1A | 0.33 | 1.41e-09 | 5.24e-07 | **0.32** | **4.03e-09** | **1.50e-06** | -2.69% |
| OPN | -0.18 | 8.07e-07 | 0.00030 | **-0.18** | **1.20e-06** | **0.00044** | -0.24% |
| IGFBP-2 | 0.21 | 1.15e-06 | 0.00043 | **0.17** | **8.42e-05** | **0.031** | -21.1% |
| CDH2 | -0.19 | 1.21e-06 | 0.00045 | **-0.17** | **9.41e-06** | **0.0035** | -10.8% |
| t-PA | -0.33 | 1.37e-06 | 0.00051 | **-0.34** | **1.27e-06** | **0.00047** | 0.81% |
| IGF2R | -0.17 | 1.40e-06 | 0.00052 | **-0.16** | **2.15e-06** | **0.00080** | -2.87% |
| HSP27 | 0.16 | 3.21e-06 | 0.0012 | **0.16** | **3.89e-06** | **0.0014** | -0.57% |
| THBS4 | -0.29 | 3.89e-06 | 0.0014 | **-0.27** | **1.29e-05** | **0.0048** | -8.80% |
| TF | -0.12 | 5.46e-06 | 0.0020 | **-0.13** | **3.09e-06** | **0.0011** | 3.87% |
| LDLreceptor | -0.21 | 6.65e-06 | 0.0025 | **-0.18** | **4.16e-05** | **0.015** | -11.8% |
| SELE | -0.18 | 7.13e-06 | 0.0027 | **-0.13** | **0.00050** | **0.19** | **-26.3%** |
| SEMA7A | -0.12 | 1.86e-05 | 0.0069 | **-0.11** | **0.00014** | **0.052** | -11.7% |
| TFPI | -0.13 | 1.87e-05 | 0.0069 | **-0.12** | **4.25e-05** | **0.016** | -4.43% |
| ITGB2 | -0.14 | 1.88e-05 | 0.0070 | **-0.13** | **6.03e-05** | **0.022** | -6.86% |
| ADGRE2 | -0.14 | 2.39e-05 | 0.0089 | **-0.14** | **6.25e-05** | **0.023** | -4.77% |
| PAI | -0.28 | 2.59e-05 | 0.0096 | **-0.24** | **0.00016** | **0.060** | -12.1% |
| HAOX1 | -0.50 | 2.99e-05 | 0.011 | **-0.45** | **0.00022** | **0.080** | -10.8% |
| SAA4 | -0.24 | 3.20e-05 | 0.012 | **-0.23** | **6.96e-05** | **0.026** | -5.05% |
| DLK-1 | -0.17 | 3.56e-05 | 0.013 | **-0.16** | **9.98e-05** | **0.037** | -5.87% |
| FCN2 | -0.14 | 4.83e-05 | 0.018 | **-0.12** | **0.00065** | **0.24** | -15.2% |
| TFF3 | 0.20 | 8.03e-05 | 0.030 | **0.20** | **0.00010** | **0.038** | -1.45% |
| COMP | -0.19 | 8.07e-05 | 0.030 | **-0.18** | **0.00015** | **0.057** | -5.07% |
| CDH5 | -0.14 | 9.85e-05 | 0.037 | **-0.13** | **0.00018** | **0.066** | -3.43% |
| SIGLEC7 | -0.11 | 9.90e-05 | 0.037 | **-0.11** | **0.00031** | **0.12** | -8.11% |
| SERPINA12 | -0.25 | 0.00010 | 0.039 | **-0.22** | **0.00074** | **0.27** | -11.4% |
| ADGRG2 | -0.16 | 0.00011 | 0.041 | **-0.17** | **4.37e-05** | **0.016** | 6.89% |
| CD97 | -0.18 | 0.00012 | 0.044 | **-0.17** | **0.00023** | **0.087** | -4.55% |
| FAM3C | 0.14 | 0.00016 | 0.058 | **0.14** | **0.00015** | **0.055** | 0.94% |
| PON3 | 0.14 | 0.00018 | 0.066 | **0.11** | **0.0034** | **1** | -**22.7%** |
| PIgR | 0.05 | 0.00018 | 0.069 | **0.054** | **0.00014** | **0.054** | 2.22% |
| MATN2 | -0.14 | 0.00020 | 0.075 | **-0.14** | **0.00028** | **0.10** | -1.71% |
| PTX3 | 0.14 | 0.00021 | 0.077 | **0.13** | **0.00033** | **0.12** | -2.05% |
| ITGA5 | -0.14 | 0.00021 | 0.079 | **-0.12** | **0.00070** | **0.26** | -10.5% |
| ICAM1 | -0.15 | 0.00021 | 0.079 | **-0.14** | **0.00051** | **0.19** | -6.63% |
| LTBR | -0.10 | 0.00028 | 0.10 | **-0.10** | **0.00036** | **0.14** | -1.16% |
| CNTN1 | -0.11 | 0.00032 | 0.12 | **-0.12** | **0.00018** | **0.068** | 5.53% |
| C1QTNF1 | -0.17 | 0.00047 | 0.17 | **-0.16** | **0.0011** | **0.41** | -8.22% |
| TYRO3 | -0.10 | 0.00051 | 0.19 | **-0.098** | **0.00099** | **0.37** | -4.61% |
| LEP | -0.19 | 0.00064 | 0.24 | **-0.074** | **0.15** | **1** | **-60.4%** |
| BCAM | -0.097 | 0.00065 | 0.24 | **-0.099** | **0.00062** | **0.23** | 1.99% |
| COL1A1 | -0.11 | 0.00069 | 0.26 | **-0.10** | **0.0011** | **0.41** | -3.85% |
| Gal-4 | 0.15 | 0.00073 | 0.27 | **0.15** | **0.00074** | **0.28** | 1.54% |
| uPA | -0.10 | 0.00090 | 0.33 | **-0.10** | **0.00087** | **0.32** | 2.02% |
| CD58 | -0.13 | 0.00091 | 0.34 | **-0.14** | **0.00085** | **0.32** | 0.79% |
| NOV | -0.15 | 0.00094 | 0.35 | **-0.14** | **0.0019** | **0.72** | -6.93% |
| CHRDL2 | -0.19 | 0.00095 | 0.35 | **-0.18** | **0.0014** | **0.54** | -3.04% |
| DPP4 | -0.12 | 0.00095 | 0.35 | **-0.12** | **0.0018** | **0.66** | -5.61% |
| ENPP7 | -0.18 | 0.00098 | 0.37 | **-0.13** | **0.015** | **1** | **-25.8%** |
| FGF-21 | -0.33 | 0.0010 | 0.38 | **-0.31** | **0.0023** | **0.85** | -6.51% |
| VCAN | -0.11 | 0.0010 | 0.39 | **-0.12** | **0.00047** | **0.17** | 8.07% |
| ALCAM | -0.083 | 0.0012 | 0.43 | **-0.080** | **0.0019** | **0.72** | -4.03% |
| IL18 | -0.12 | 0.0012 | 0.44 | **-0.11** | **0.0022** | **0.81** | -4.40% |
| FCGR3B | -0.11 | 0.0013 | 0.48 | **-0.10** | **0.0033** | **1** | -7.24% |
| NT-proBNP | 0.36 | 0.0013 | 0.50 | **0.33** | **0.0030** | **1** | -7.09% |
| FSTL3 | -0.096 | 0.0014 | 0.53 | **-0.088** | **0.0034** | **1** | -8.44% |
| THOP1 | -0.12 | 0.0017 | 0.62 | **-0.11** | **0.0038** | **1** | -8.33% |
| CD300LG | -0.13 | 0.0019 | 0.69 | **-0.13** | **0.0020** | **0.76** | 0.11% |
| AP-N | -0.074 | 0.0021 | 0.78 | **-0.073** | **0.0027** | **0.99** | -1.20% |
| TNFSF13B | -0.095 | 0.0023 | 0.87 | **-0.094** | **0.0030** | **1** | -1.33% |
| TIE1 | -0.081 | 0.0026 | 0.97 | **-0.076** | **0.0048** | **1** | -6.81% |
| CNTN4 | -0.088 | 0.0026 | 0.97 | **-0.086** | **0.0036** | **1** | -2.48% |
| MERTK | -0.096 | 0.0026 | 0.97 | **-0.098** | **0.0025** | **0.92** | 1.77% |
| CHI3L1 | -0.22 | 0.0031 | 1 | **-0.18** | **0.013** | **1** | -17.6% |
| FETUB | -0.15 | 0.0034 | 1 | **-0.14** | **0.0042** | **1** | -2.18% |
| PGF | -0.093 | 0.0039 | 1 | **-0.092** | **0.0046** | **1** | -1.56% |
| OMD | -0.13 | 0.0040 | 1 | **-0.14** | **0.0031** | **1** | 4.59% |
| AOC3 | -0.097 | 0.0040 | 1 | **-0.093** | **0.0061** | **1** | -4.43% |
| B4GALT1 | -0.16 | 0.0048 | 1 | **-0.16** | **0.0053** | **1** | -1.22% |
| SCGB3A1 | -0.15 | 0.0051 | 1 | **-0.17** | **0.0015** | **0.56** | 14.7% |
| CA6 | -0.10 | 0.0052 | 1 | **-0.10** | **0.0046** | **1** | 4.14% |
| SRC | 0.14 | 0.0052 | 1 | **0.14** | **0.0048** | **1** | 1.17% |
| SELL | -0.10 | 0.0052 | 1 | **-0.091** | **0.0098** | **1** | -7.32% |
| CD93 | -0.073 | 0.0054 | 1 | **-0.072** | **0.0068** | **1** | -1.63% |
| Notch3 | -0.11 | 0.0056 | 1 | **-0.12** | **0.0038** | **1** | 6.26% |
| HO-1 | -0.097 | 0.0057 | 1 | **-0.10** | **0.0037** | **1** | 6.19% |
| CCL15 | 0.087 | 0.0062 | 1 | **0.082** | **0.012** | **1** | -5.52% |
| CD1C | -0.087 | 0.0068 | 1 | **-0.087** | **0.0072** | **1** | 0.50% |
| IL-1RT2 | -0.082 | 0.0070 | 1 | **-0.064** | **0.032** | **1** | **-21.7%** |
| CDHR5 | -0.083 | 0.0071 | 1 | **-0.060** | **0.051** | **1** | **-27.9%** |
| ITGB1 | -0.087 | 0.0072 | 1 | **-0.083** | **0.010** | **1** | -4.47% |
| CD99L2 | -0.084 | 0.0073 | 1 | **-0.079** | **0.012** | **1** | -5.69% |
| LRP11 | -0.091 | 0.0073 | 1 | **-0.086** | **0.011** | **1** | -5.49% |
| TIMD4 | -0.11 | 0.0077 | 1 | **-0.099** | **0.020** | **1** | -13.2% |
| CD79B | -0.081 | 0.0079 | 1 | **-0.077** | **0.011** | **1** | -4.56% |
| BLMhydrolase | -0.11 | 0.0080 | 1 | **-0.11** | **0.0093** | **1** | -1.61% |
| ROR1 | -0.088 | 0.0084 | 1 | **-0.096** | **0.0045** | **1** | 9.01% |
| CDON | -0.097 | 0.0085 | 1 | **-0.095** | **0.0099** | **1** | -1.75% |
| CRELD2 | -0.18 | 0.0086 | 1 | **-0.17** | **0.013** | **1** | -5.79% |
| OSMR | -0.070 | 0.0088 | 1 | **-0.068** | **0.011** | **1** | -2.43% |
| SCF | 0.084 | 0.0097 | 1 | **0.067** | **0.036** | **1** | -19.9% |
| FCRL1 | -0.087 | 0.011 | 1 | **-0.072** | **0.038** | **1** | -17.6% |
| ROBO1 | -0.11 | 0.011 | 1 | **-0.10** | **0.013** | **1** | -2.32% |
| ENG | -0.10 | 0.011 | 1 | **-0.10** | **0.011** | **1** | 0.76% |
| TGFBI | -0.11 | 0.013 | 1 | **-0.096** | **0.024** | **1** | -9.59% |
| CLEC14A | -0.085 | 0.013 | 1 | **-0.088** | **0.011** | **1** | 4.07% |
| CCDC80 | -0.094 | 0.013 | 1 | **-0.092** | **0.016** | **1** | -1.38% |
| ACE2 | -0.12 | 0.014 | 1 | **-0.097** | **0.059** | **1** | **-22.3%** |
| IL7R | -0.15 | 0.016 | 1 | **-0.15** | **0.015** | **1** | 1.85% |
| TPP1 | -0.11 | 0.017 | 1 | **-0.099** | **0.031** | **1** | -9.40% |
| SUMF2 | -0.13 | 0.017 | 1 | **-0.12** | **0.037** | **1** | -12.8% |
| CTSZ | -0.074 | 0.017 | 1 | **-0.062** | **0.044** | **1** | -15.7% |
| SEMA3F | -0.075 | 0.017 | 1 | **-0.069** | **0.028** | **1** | -8.13% |
| KIM1 | -0.12 | 0.017 | 1 | **-0.096** | **0.056** | **1** | -18.8% |
| Gal-3 | 0.074 | 0.017 | 1 | **0.068** | **0.032** | **1** | -8.96% |
| IL13RA1 | -0.13 | 0.018 | 1 | **-0.13** | **0.022** | **1** | -3.44% |
| EFEMP1 | -0.10 | 0.018 | 1 | **-0.098** | **0.026** | **1** | -5.87% |
| FUCA1 | -0.088 | 0.018 | 1 | **-0.014** | **0.71** | **1** | **-84.2%** |
| SOD2 | -0.064 | 0.020 | 1 | **-0.062** | **0.023** | **1** | -2.31% |
| IGFBP-7 | -0.073 | 0.020 | 1 | **-0.065** | **0.039** | **1** | -10.3% |
| LILRA5 | -0.073 | 0.021 | 1 | **-0.059** | **0.064** | **1** | **-20.2%** |
| TIE2 | -0.061 | 0.021 | 1 | **-0.059** | **0.029** | **1** | -4.31% |
| ACAN | -0.068 | 0.022 | 1 | **-0.069** | **0.021** | **1** | 1.27% |
| EGFR | -0.051 | 0.022 | 1 | **-0.050** | **0.024** | **1** | -1.19% |
| THBS2 | -0.041 | 0.022 | 1 | **-0.037** | **0.041** | **1** | -9.63% |
| CTSO | -0.089 | 0.024 | 1 | **-0.080** | **0.041** | **1** | -10.0% |
| PAM | -0.087 | 0.024 | 1 | **-0.083** | **0.031** | **1** | -4.89% |
| PDGFRB | 0.066 | 0.026 | 1 | **-0.058** | **0.051** | **1** | -12.2% |
| TNFRSF11A | 0.070 | 0.027 | 1 | **0.081** | **0.012** | **1** | 15.9% |
| GAS6 | -0.098 | 0.028 | 1 | **-0.091** | **0.040** | **1** | -6.59% |
| OPG | -0.073 | 0.029 | 1 | **-0.066** | **0.049** | **1** | -9.43% |
| CTSD | -0.085 | 0.029 | 1 | **-0.069** | **0.072** | **1** | -18.0% |
| NOTCH1 | -0.078 | 0.030 | 1 | **-0.077** | **0.032** | **1** | -0.94% |
| LAMA4 | -0.10 | 0.032 | 1 | **-0.097** | **0.040** | **1** | -4.56% |
| SIRPB1 | -0.086 | 0.032 | 1 | **-0.083** | **0.041** | **1** | -3.44% |
| EPHB4 | -0.062 | 0.032 | 1 | **-0.063** | **0.032** | **1** | 1.44% |
| LILRB1 | -0.072 | 0.033 | 1 | **-0.066** | **0.052** | **1** | -8.88% |
| SPINT1 | -0.095 | 0.033 | 1 | **-0.093** | **0.038** | **1** | -2.56% |
| HAVCR2 | -0.077 | 0.033 | 1 | **-0.071** | **0.052** | **1** | -8.59% |
| TM | -0.055 | 0.034 | 1 | **-0.052** | **0.045** | **1** | -4.77% |
| CCL21 | -0.081 | 0.034 | 1 | **-0.075** | **0.053** | **1** | -7.91% |
| PLC | -0.051 | 0.034 | 1 | **-0.045** | **0.060** | **1** | -11.6% |
| CLSTN2 | -0.078 | 0.035 | 1 | **-0.067** | **0.072** | **1** | -14.0% |
| PTPRS | -0.071 | 0.035 | 1 | **-0.070** | **0.037** | **1** | -1.00% |
| DDC | -0.12 | 0.035 | 1 | **-0.13** | **0.028** | **1** | 6.89% |
| CEACAM8 | 0.11 | 0.036 | 1 | **0.12** | **0.032** | **1** | 2.92% |
| ICAM-2 | -0.052 | 0.037 | 1 | **-0.051** | **0.050** | **1** | -2.90% |
| CTRC | 0.13 | 0.038 | 1 | **0.12** | **0.070** | **1** | -11.5% |
| CTSL1 | -0.055 | 0.039 | 1 | **-0.053** | **0.047** | **1** | -2.67% |
| FGF-23 | -0.082 | 0.039 | 1 | **-0.075** | **0.063** | **1** | -9.34% |
| GUSB | -0.094 | 0.041 | 1 | **-0.067** | **0.14** | **1** | **-29.5%** |
| LILRB2 | -0.071 | 0.041 | 1 | **-0.061** | **0.077** | **1** | -13.1% |
| vWF | -0.20 | 0.043 | 1 | **-0.20** | **0.044** | **1** | -0.48% |
| IL-18BP | -0.056 | 0.045 | 1 | **-0.050** | **0.078** | **1** | -11.1% |
| IGFBP3 | -0.071 | 0.046 | 1 | **-0.072** | **0.047** | **1** | 1.39% |
| XG | -0.092 | 0.047 | 1 | **-0.087** | **0.062** | **1** | -5.22% |
| NOMO1 | -0.078 | 0.049 | 1 | **-0.068** | **0.085** | **1** | -13.1% |
| PEAR1 | -0.079 | 0.049 | 1 | **-0.077** | **0.055** | **1** | -2.14% |
| SERPINA7 | -0.073 | 0.050 | 1 | **-0.066** | **0.077** | **1** | -10.4% |

Supplementary Table 9 showing a combined analysis of RAMP and S3WP-T2D. Results are shown for an unadjusted analysis and following adjustment for BMI change. Any Bonferroni adjusted P value less than 0.05 is highlighted in red. Percentage attenuation of the estimate following BMI adjustment is also shown.

Supplementary Table 10- Comparison of Longitudinal Olink analysis between males and females

Males (n=54)	Females (n=44)

| Protein | Estimate | P value | Adjusted P Value | Estimate | P value | Adjusted P Value |
| --- | --- | --- | --- | --- | --- | --- |
| REG4 | **0.66** | **4.66E-15** | **1.73E-12** | **0.84** | **1.87E-20** | **6.96E-18** |
| GDF-15 | **0.70** | **2.02E-16** | **7.51E-14** | **0.93** | **1.10E-17** | **4.09E-15** |
| Ep-CAM | **-1.13** | **2.72E-15** | **1.01E-12** | **-1.17** | **8.88E-14** | **3.30E-11** |
| SPINK1 | **0.49** | **3.20E-12** | **1.19E-09** | **0.64** | **1.48E-16** | **5.51E-14** |
| REG1A | **0.32** | **3.03E-05** | **0.011** | **0.48** | **3.76E-14** | **1.40E-11** |
| LDLreceptor | **-0.27** | **4.84E-07** | **0.00018** | **-0.26** | **5.06E-07** | **0.00019** |
| IGFBP-2 | **0.26** | **6.50E-08** | **2.42E-05** | 0.16 | 0.00098 | 0.36 |
| t-PA | **-0.41** | **8.74E-08** | **3.25E-05** | -0.29 | 0.0018 | 0.67 |
| CDH2 | **-0.22** | **9.63E-08** | **3.58E-05** | -0.12 | 0.0040 | 1 |
| SEMA7A | **-0.14** | **4.42E-05** | **0.016** | **-0.14** | **1.19E-05** | **0.0044** |
| Gal-4 | **0.21** | **2.26E-05** | **0.0084** | **0.23** | **5.88E-05** | **0.022** |
| SAA4 | **-0.35** | **2.57E-07** | **9.56E-05** | -0.14 | 0.0050 | 1 |
| TFF3 | **0.25** | **3.66E-05** | **0.014** | **0.19** | **3.58E-07** | **0.00013** |
| OPN | **-0.17** | **3.95E-05** | **0.015** | **-0.16** | **5.96E-05** | **0.022** |
| LEP | **-0.28** | **4.84E-06** | **0.0018** | -0.23 | 0.00079 | 0.30 |
| FCN2 | **-0.20** | **8.87E-07** | **0.00033** | -0.095 | 0.0022 | 0.81 |
| ADGRE2 | **-0.16** | **2.31E-06** | **0.00086** | -0.12 | 0.0019 | 0.72 |
| DLK-1 | **-0.17** | **6.45E-05** | **0.024** | -0.15 | 0.00014 | 0.053 |
| IGF2R | **-0.18** | **1.32E-05** | **0.0049** | -0.12 | 0.0012 | 0.46 |
| COMP | **-0.24** | **4.91E-06** | **0.0018** | -0.16 | 0.0031 | 1 |
| SELE | **-0.18** | **1.69E-05** | **0.0063** | -0.12 | 0.0023 | 0.84 |
| CDH5 | -0.12 | 0.00048 | 0.18 | **-0.14** | **5.90E-05** | **0.022** |
| ICAM1 | **-0.18** | **4.64E-05** | **0.017** | -0.11 | 0.0037 | 1 |
| ITGB2 | **-0.15** | **0.00011** | **0.040** | -0.12 | 0.0013 | 0.50 |
| TF | **-0.13** | **6.75E-06** | **0.0025** | -0.065 | 0.019 | 1 |
| CNTN1 | -0.11 | 0.00087 | 0.32 | -0.12 | 0.00017 | 0.061 |
| NOV | **-0.19** | **3.26E-05** | **0.012** | -0.15 | 0.0057 | 1 |
| ADGRG2 | **-0.17** | **1.12E-05** | **0.0042** | -0.11 | 0.018 | 1 |
| FETUB | **-0.28** | **1.21E-06** | **0.00045** | -0.085 | 0.12 | 1 |
| SIGLEC7 | **-0.15** | **1.96E-07** | **7.29E-05** | -0.051 | 0.13 | 1 |
| FAM3C | 0.095 | 0.022 | 1 | **0.20** | **6.33E-07** | **0.00024** |
| CD97 | -0.18 | 0.00028 | 0.10 | -0.14 | 0.0028 | 1 |
| uPA | -0.12 | 0.00047 | 0.17 | -0.098 | 0.0028 | 1 |
| MERTK | -0.11 | 0.0021 | 0.79 | -0.13 | 0.00025 | 0.093 |
| TIE1 | **-0.13** | **2.59E-05** | **0.0096** | -0.057 | 0.036 | 1 |
| CHRDL2 | **-0.24** | **1.14E-06** | **0.00042** | -0.15 | 0.040 | 1 |
| HAOX1 | -0.37 | 0.013 | 1 | **-0.60** | **8.20E-06** | **0.0031** |
| FCGR3B | **-0.18** | **2.47E-06** | **0.00092** | -0.053 | 0.14 | 1 |
| DPP4 | **-0.17** | **2.92E-05** | **0.011** | -0.072 | 0.038 | 1 |
| PON3 | 0.12 | 0.00090 | 0.33 | 0.13 | 0.0020 | 0.73 |
| AP-N | -0.077 | 0.0023 | 0.87 | -0.088 | 0.00063 | 0.23 |
| LTBR | -0.091 | 0.0030 | 1 | -0.11 | 0.00048 | 0.18 |
| TYRO3 | **-0.12** | **9.68E-06** | **0.0036** | -0.071 | 0.042 | 1 |
| SERPINA12 | -0.23 | 0.00090 | 0.34 | -0.23 | 0.0029 | 1 |
| COL1A1 | **-0.13** | **4.80E-05** | **0.018** | -0.086 | 0.029 | 1 |
| ENPP7 | **-0.25** | **7.78E-06** | **0.0029** | -0.093 | 0.11 | 1 |
| OMD | -0.19 | 0.00014 | 0.051 | -0.14 | 0.019 | 1 |
| Notch3 | -0.13 | 0.0021 | 0.80 | -0.12 | 0.0014 | 0.52 |
| VCAN | -0.14 | 0.00021 | 0.076 | -0.081 | 0.026 | 1 |
| PAI | -0.23 | 0.0012 | 0.46 | -0.18 | 0.0020 | 0.75 |
| AOC3 | **-0.14** | **6.90E-05** | **0.026** | -0.070 | 0.043 | 1 |
| BCAM | -0.099 | 0.00083 | 0.31 | -0.092 | 0.0059 | 1 |
| CCL15 | 0.085 | 0.015 | 1 | **0.14** | **8.06E-05** | **0.030** |
| CD300LG | -0.15 | 0.00076 | 0.28 | -0.12 | 0.0061 | 1 |
| SELL | -0.14 | 0.00021 | 0.076 | -0.079 | 0.031 | 1 |
| THOP1 | **-0.16** | **8.44E-05** | **0.031** | -0.080 | 0.052 | 1 |
| THBS4 | **-0.32** | **3.94E-06** | **0.0015** | -0.21 | 0.079 | 1 |
| CNTN4 | -0.12 | 0.00076 | 0.28 | -0.081 | 0.0085 | 1 |
| IL18 | -0.12 | 0.00097 | 0.36 | -0.10 | 0.010 | 1 |
| TFPI | **-0.12** | **0.00011** | **0.040** | -0.066 | 0.051 | 1 |
| ALCAM | -0.084 | 0.0019 | 0.72 | -0.070 | 0.0074 | 1 |
| CD93 | -0.081 | 0.0046 | 1 | -0.082 | 0.0012 | 0.46 |
| PIgR | 0.050 | 0.0014 | 0.53 | 0.037 | 0.014 | 1 |
| FUCA1 | -0.14 | 0.00088 | 0.33 | -0.089 | 0.027 | 1 |
| TIMD4 | **-0.20** | **6.37E-05** | **0.024** | -0.049 | 0.19 | 1 |
| CDON | -0.12 | 0.0028 | 1 | -0.10 | 0.011 | 1 |
| PGF | -0.12 | 0.00096 | 0.36 | -0.069 | 0.040 | 1 |
| CD58 | -0.12 | 0.0048 | 1 | -0.11 | 0.0091 | 1 |

Supplementary Table 10 showing the comparison between males and females for the 68 significantly changed proteins in the Longitudinal Olink analysis. Significantly adjusted p values after bonferroni correction (adj p<0.05) are highlighted in bold.

Link to Supplementary excel file containing results for all proteins irrespective of significance in each of the three studies:

<https://dmail-my.sharepoint.com/:x:/g/personal/bwconnolly_dundee_ac_uk/EcEc3jZ08l5MvrRMXZnxf_0B0IBtwcTZWQLu-JCgw2_eCA?e=jZ4asB>
